# A modeling-based approach to optimize COVID-19 vaccine dosing schedules for improved protection

**DOI:** 10.1101/2022.09.14.22279959

**Authors:** Prashant Dogra, Carmine Schiavone, Zhihui Wang, Javier Ruiz-Ramírez, Sergio Caserta, Daniela I. Staquicini, Christopher Markosian, Jin Wang, H. Dirk Sostman, Renata Pasqualini, Wadih Arap, Vittorio Cristini

## Abstract

While the development of different vaccines has slowed the dissemination of SARS-CoV-2, the occurrence of breakthrough infections continues to fuel the pandemic. As a strategy to secure at least partial protection, with a single dose of a given COVID-19 vaccine to maximum possible fraction of the population, *delayed* administration of subsequent doses (or boosters) has been implemented in many countries. However, waning immunity and emergence of new variants of SARS-CoV-2 suggest that such measures may jeopardize the attainment of herd immunity due to intermittent lapses in protection. Optimizing vaccine dosing schedules could thus make the difference between periodic occurrence of breakthrough infections or effective control of the pandemic. To this end, we have developed a mechanistic mathematical model of adaptive immune response to vaccines and demonstrated its applicability to COVID-19 mRNA vaccines as a proof-of-concept for future outbreaks. The model was thoroughly calibrated against multiple clinical datasets involving immune response to SARS-CoV-2 infection and mRNA vaccines in healthy and immunocompromised subjects (cancer patients undergoing therapy); the model showed robust clinical validation by accurately predicting neutralizing antibody kinetics, a correlate of vaccine-induced protection, in response to multiple doses of mRNA vaccines. Importantly, we estimated population vulnerability to breakthrough infections and predicted tailored vaccination dosing schedules to maximize protection and thus minimize breakthrough infections, based on the immune status of a sub-population. We have identified a critical waiting window for cancer patients (or, immunocompromised subjects) to allow recovery of the immune system (particularly CD4+ T-cells) for effective differentiation of B-cells to produce neutralizing antibodies and thus achieve optimal vaccine efficacy against variants of concern, especially between the first and second doses. Also, we have obtained optimized dosing schedules for subsequent doses in healthy and immunocompromised subjects, which vary from the CDC-recommended schedules, to minimize breakthrough infections. The developed modeling tool is based on generalized adaptive immune response to antigens and can thus be leveraged to guide vaccine dosing schedules during future outbreaks.

## 1. Introduction

Since December 2019, the COVID-19 pandemic caused by SARS-CoV-2 has afflicted more than 655 million individuals and caused more than 6.67 million deaths worldwide [1]. Global vaccination programs along with public health measures such as social distancing and masking are anticipated to be the most effective approaches to attain herd immunity and curb the pandemic [2, 3]. Herd immunity represents a scenario where a virus cannot spread due to a dearth of susceptible hosts and can be achieved through natural infection and/or vaccination of the population. In December 2020, the first COVID-19 vaccine obtained Emergency Use Authorization from the United States Food and Drug Administration (FDA), and as of December 2022, 50 vaccines have obtained regulatory approval in at least one country [4]. As a result, over 63.4% of the world population is fully vaccinated and ∼69% of the population has received at least a single dose of a COVID-19 vaccine. However, due to the inequitable allocation of vaccines, only ∼26% of the people in low-income countries have received at least a single dose [5, 6], which cans facilitate the emergence of new variants of SARS-CoV-2 and thus resurgence of the pandemic.

According to a meta-analysis, seroconversion rates related to the development of neutralizing antibodies in the sera of individuals doubly vaccinated with COVID-19 vaccines have been found to be dependent on patient immunological health status; seroconversion positivity in immunocompetent individuals can be up to 99%, while in immunosuppressed patients the efficacy of vaccination varies for different diseases (e.g., solid tumors ∼92%, immune-mediated inflammatory diseases ∼78%, hematological cancers ∼64%, and organ transplant recipients ∼27%) [7, 8]. Due to limited protection, immunocompromised individuals are more vulnerable to infection and are at a higher risk of developing severe or lethal COVID-19. Thus, immunizing the majority of the population is a means to additionally protect individuals who are susceptible or unable to receive a vaccine.

However, the emergence of breakthrough infections in previously infected or vaccinated individuals is a major challenge. The key biological reasons for breakthrough infections are attributed to: (i) waning immunity over time, and (ii) emergence of mutant variants of SARS-CoV-2, referred to as variants of concern (VOCs) [9, 10]. Depending on demographics and the type of vaccine administered, the humoral response (i.e., neutralizing antibodies) against SARS-CoV-2 has been found to be substantially reduced within about six months after two-dose vaccination [11-13]. Thus, vaccines with an initial effectiveness of 90% are only ∼30-70% effective after six months [14-16]. Further, coronaviruses tend to have high genetic diversity due to their large genome size (26.4 – 31.7 kb), high mutation rate caused by a low-fidelity viral polymerase (∼10^−4^ substitutions per site per year), and high recombination frequency (up to 25% for the entire genome *in vivo*) [17]. As a result of selection pressure imposed by neutralizing antibodies on viral surface proteins, particularly the receptor binding domain (RBD) and the N-terminal domain (NTD) of the spike protein, which are the targets of most of the COVID-19 vaccine-induced neutralizing antibodies, SARS-CoV-2 show clusters of mutations as documented in the genomes of VOCs [18]. Mutations that confer greater fitness such as increased transmission rates and improved antibody escape are positively selected, leading to antigenic drift that makes the vaccination-induced neutralizing antibodies partially ineffective against the mutant strains [17]. This predisposes the vaccinated or previously infected individuals to breakthrough infections [19] (though the severity of symptoms tends to be milder) [20].

Currently, additional (booster) doses of COVID-19 vaccines are being used to reinforce protection and minimize breakthrough infections [21-24]. Boosters are being administered to fully vaccinated individuals since ∼June 2021, except in low-income countries [25], and prioritized for high-risk populations such as the elderly and immunocompromised patients [26]. According to the Centers for Disease Control and Prevention (CDC), a two-dose schedule (3- to 8-week gap) followed by a third dose (5-month gap) of mRNA vaccine (Pfizer-BioNTech or Moderna) is recommended for immunocompetent adults, while a three-dose schedule (3- to 4-week gap between doses 1, 2, and 3) followed by a fourth dose (12-week gap) is recommended for immunocompromised adults [27]. These scheduling recommendations are based on clinical trials, which are generally limited to healthy volunteers, thereby may require optimization, especially for special populations, to achieve better protection at the population scale. A mathematical modeling approach, which is data-driven and based on first principles of physiology, immunology, and biophysics can be a valuable tool to simulate population-scale heterogeneity in immune health status and immune response to vaccines, thereby supporting rational design of dosing schedules. In addition, given the disparities in global vaccine allocation, optimization of dosing schedule to extend the gaps between doses with no major effect on efficacy could allow for improved distribution of vaccines to countries without the capacity to provide for themselves, reduce costs, and promote vaccine compliance, thereby benefiting the overall population, but especially patients in critical care.

Using a mathematical modeling approach, we identified optimal vaccine dosing schedules of mRNA COVID-19 vaccines for immunocompetent and immunocompromised individuals to minimize breakthrough infections at the population scale. Clinical evidence that demonstrates acceptable vaccine effectiveness, despite delayed follow-up doses, sets the premise for our theoretical investigation [28-30]. Previous mathematical models that have been developed to identify optimal vaccine allocation and dosing schedules to minimize hospitalizations and deaths due to COVID-19 are primarily age-structured compartmental models, based on epidemiological principles (e.g., susceptible, exposed, infectious, and removed (SEIR) models), which focus on the transmission of the virus under different vaccination scenarios and the analysis of strategies to reduce the rate of infection [31-37]. These models, however, lack mechanistic details relevant to virus-host interaction, the immune response to vaccines, and the time-dependent variation in vaccine efficacy due to inter-individual variability, vaccine efficacy against VOCs, and other biological/physiological factors. To this end, as an adaptation of our previous mechanistic models of complex biological systems [38-43], we have developed a mathematical model that accurately simulates the adaptive immune response to COVID-19 vaccines at the individual scale. The model was calibrated and validated with clinical data for mRNA-based COVID-19 vaccines to conduct analysis in virtual cohorts comprising immunocompetent and immunocompromised digital twins (virtual cancer patients undergoing chemotherapy and/or immunotherapy). The model identified optimal schedules for vaccination doses that minimize vulnerability to breakthrough infections, especially against VOCs (specifically Omicron), while retaining vaccine efficacy above the protection threshold in populations with different health statuses.

## 2. Methods

### 2.1 Model development

Based on our previous mathematical modeling of the immune response to SARS-CoV-2 infection [44], we developed a model of the adaptive immune response to COVID-19 vaccines. As shown in **Figure 1**, the model incorporates key biological processes that are relevant to antigen presentation at the site of vaccination (i.e., muscle), the development of adaptive immune responses in the lymphoid tissue, and protection against infection in the respiratory tract. The model was formulated as a system of ordinary differential equations (ODEs, **Equations 1-17**), which describe the kinetics of key immune response variables following vaccination or infection. The equations were solved numerically as an initial value problem in MATLAB R2018a. While some of the model parameters were known a priori (**Table 1**), the remainder were estimated by non-linear least squares fitting of the model to multiple clinical datasets obtained from the literature [45-47]. The model was then used to simulate the immune response to mRNA-based COVID-19 vaccines in healthy and immunocompromised populations and was implemented to identify optimal vaccine dosing schedules to minimize breakthrough infections. The model equations are described in detail below.

**Table 1.**
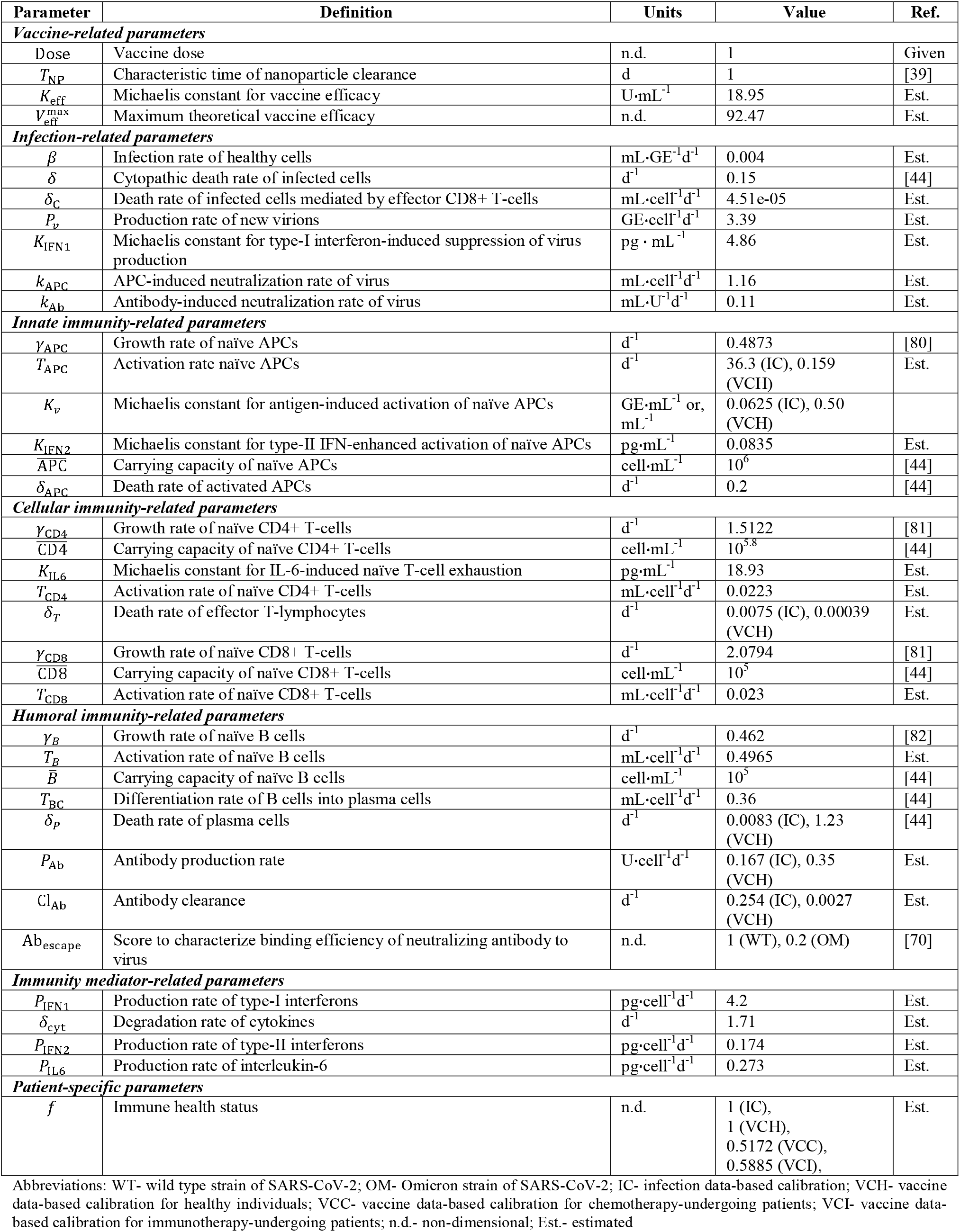
List of model parameters.

**Figure 1.**
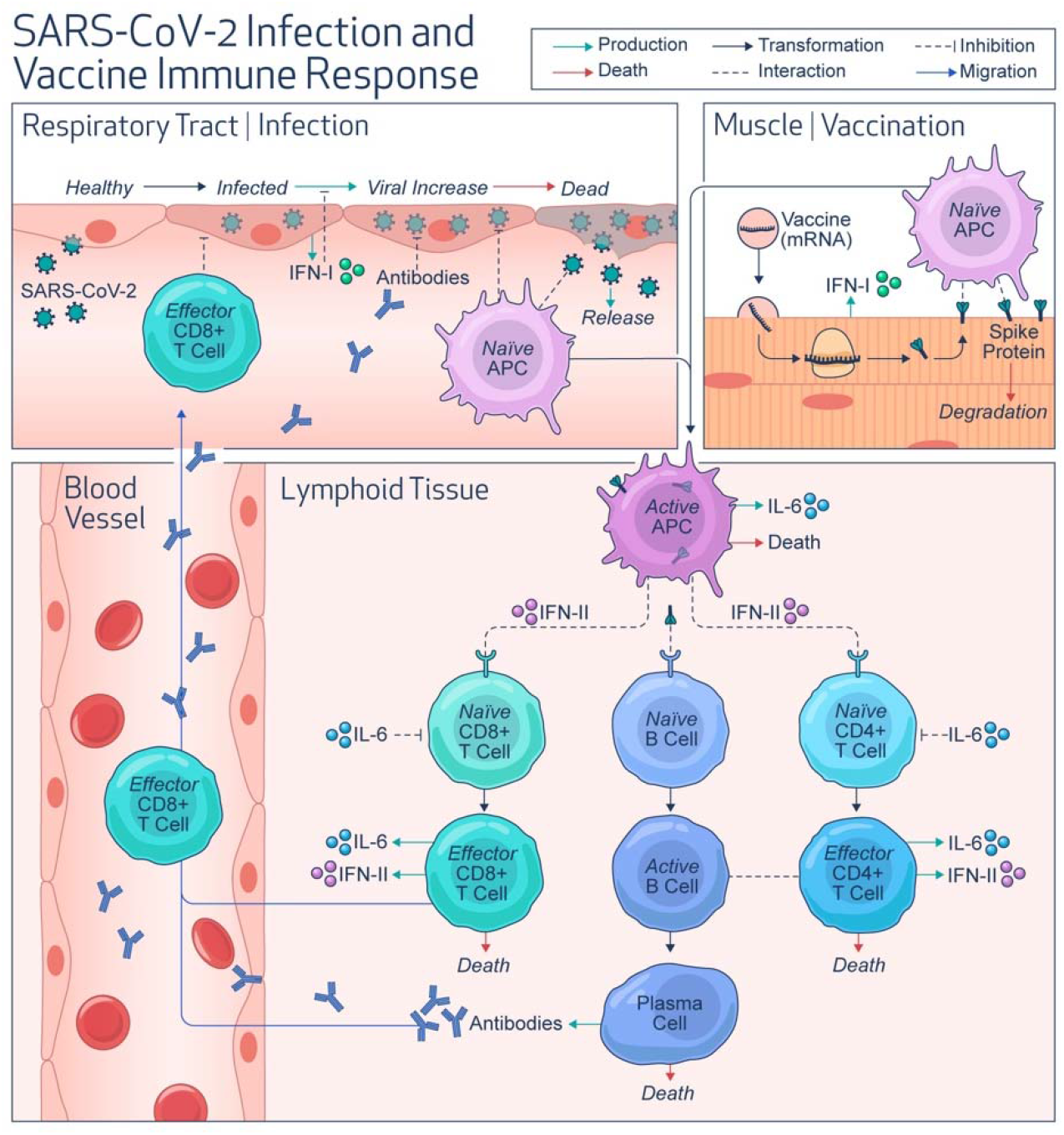
Model schematic. Diagram shows key variables and system interactions incorporated into the mathematical model. Upon respiratory tract infection by SARS-CoV-2 or intramuscular administration of mRNA vaccines, antigen presenting cells (e.g., macrophages) engage the adaptive immune system to produce antibodies and activate T-lymphocytes to build immunity against infection. Cytokines secreted by infected cells (e.g., IFN-I) and immune cells (e.g., IFN-II, IL-6) in the process have modulatory effects on the immune system. Abbreviations: IFN-I, type-I interferon; IFN-II, type-II interferon; IL-6, interleukin 6.

At the site of vaccination, nanoparticles carrying the mRNA of SARS-CoV-2 spike protein are endocytosed into myocytes, leading to the translation and expression of spike protein on myocytes [48]. Given that the timescale of drug delivery (intramuscular injection) and mRNA translation is much shorter (< 1 hour) [49] than that of the vaccine-induced immune response (days to weeks) [50], we assumed that the variable *C*_*a*_(*t*) represents the concentration of vaccine-induced spike protein in the muscle cells that can trigger the immune response via antigen-presenting cells (APCs).

#### Concentration kinetics of the exogenously administered antigen (via vaccine) in muscle cells (*C*_*a*_ (*t*))

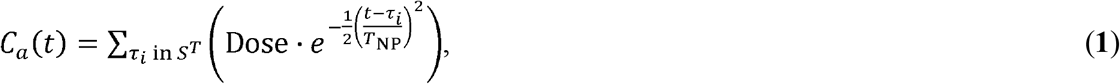

where Dose indicates the dimensionless dose of the antigen administered via the vaccine. The concentration of the spike protein *C*_*a*_(*t*) is described by the sum of Gaussians centered at τ_*i*_, which represents the day on which a vaccine dose is injected out of the set of doses indicated by *S*^*T*^. *T*_NP_ is the characteristic time of clearance of the antigen-carrying nanoparticle (NP) from the body [39], estimated based on NP diameter of 100 nm for mRNA vaccines [51].

The population of naïve (or immature) APCs is maintained through continuous regeneration and presumably maintained at a steady state. Thus, we used a logistic growth term to include this contribution, where *γ*_APC_ is the exponential growth rate, and 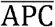 is the carrying capacity of the APC population. Naïve APCs at the site of expression of spike proteins recognize, process, and present the antigen via major histocompatibility complex (MHC) during differentiation into activated APC (APC*) at a rate *T*_APC_ as they migrate towards the lymphoid tissue. The APC activation process is proportional to the antigen load (Ag(*t*)), which can be derived either from the vaccine or natural infection and is either equal to *C*_*a*_(*t*) or the viral load *V*(*t*) in the case of vaccination or infection, respectively, with *K*_*ν*_ being the Michaelis constant for antigen-induced activation of naïve APCs.

#### Equation for the naïve APC density at the site of vaccination or natural infection (APC(*t*))

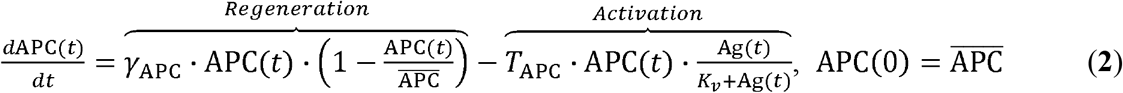

Activated APCs are primarily responsible for the induction of the adaptive immune response, and their population is determined by the activation of naïve APCs, which we discussed in Eq. 2, and a death term determined by the death rate constant, *δ*_APC_ of activated APCs.

#### Equation for the activated APC density (APC*(*t*))

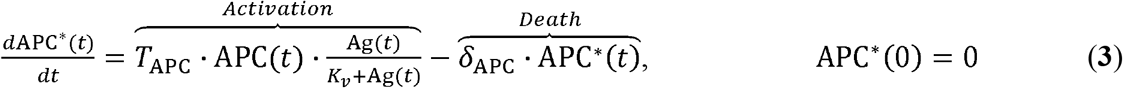

Activated APCs migrate from the site of vaccination or natural infection to the lymphoid tissue to interact with naïve T-cells (CD8+ or CD4+) and transform them into their active or effector forms. Alternatively, naïve B-cells are activated by the binding of soluble antigens, which however in the current model is replaced by binding to active APCs, given that the density of active APCs is dependent on antigen load in the body. For the naïve cells, population density is determined by cell regeneration and cell activation, where we used a logistic growth term with *γ*_CD4_, *γ*_CD8_, and *γ*_B_ as the growth rates of naïve forms of CD4+ T-cells, CD8+ T-cells, and B cells, respectively; 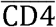, 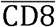, and 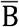 are the carrying capacities of the corresponding cell the product of active APC density and the corresponding naïve cell density; *T*_CD4_,*T*_CD8_ and *T*_B_ populations, respectively. The activation term has second-order kinetics and is proportional to are the activation rates of lymphocytes indicated by the subscript. Activation of T-cells is amplified by the presence of type-II interferons (IFN2(*t*)) secreted by activated T-cells [52], with possible saturation effects. Thus, we used a Michaelis-Menten term to model this process in which *K*_IFN2_ is the Michaelis constant of type-II interferon effects.

Of note, in our model we have included a dimensionless coefficient *f* ∈ [0, 1] that represents an immunosuppression factor to modulate the carrying capacity (i.e., homeostasis levels) of the naïve immune cell population to model immunocompromised subjects, such that *f* = 1 in healthy individuals, and *f* < 1 in immunocompromised patients. Note that since our model is calibrated for patients who are immunocompromised due to anticancer therapy, therefore immunosuppression in our model is characterized by T- and B-cell immunodeficiency, which is one of the important immunological effects observed due to disruption of hematopoiesis leading to myelosuppression in patients undergoing anticancer therapy [53-57]. Also, in the case of naïve CD4+ and CD8+ T-cells we have included the ability of interleukin-6 (IL-6) to cause T-cell exhaustion [58] by including an additional term that limits the carrying capacity of these cells. This term uses the concentration of IL-6 in a Michaelis-Menten function, where, *K*_IL6_ is the Michaelis constant for IL-6 effects.

#### Equation for the naïve CD4+ T-cell density (CD4(*t*))

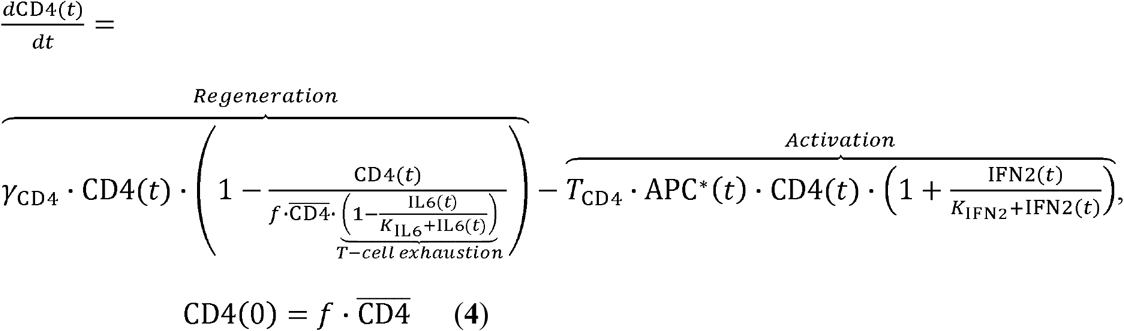

#### Equation for the effector CD4+ T-cell density (CD4*(*t*))

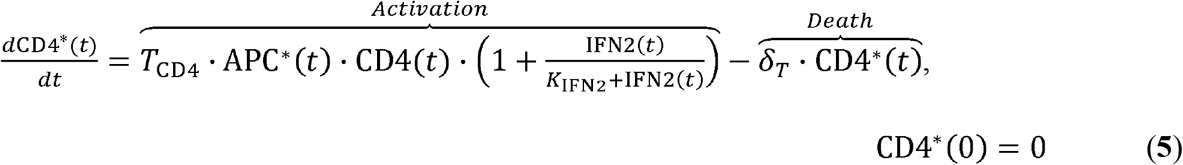

where *δ*_*T*_ is the death rate of effector T-cells.

#### Equation for the naïve CD8+ T-cell density (CD8(*t*))

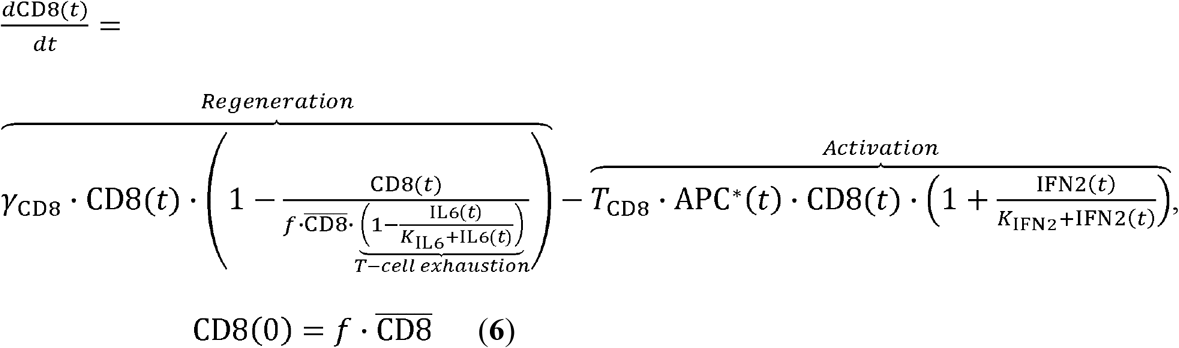

#### Equation for the effector CD8+ T-cell density (CD8*(*t*))

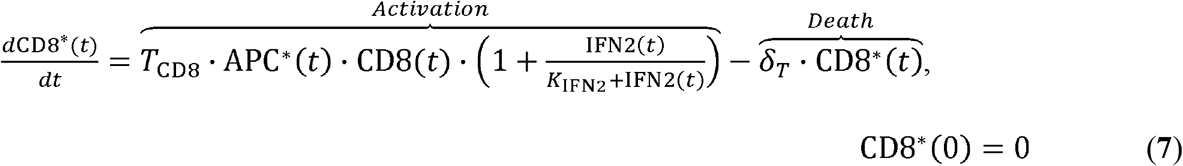

#### Equation for the naïve B cell density (*B*(*t*))

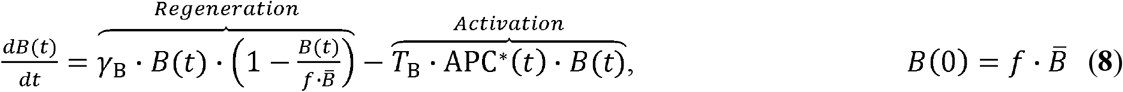

where *T*_B_ is the transition rate of naïve B cells into their activated form.

Of note, the activated B cells differentiate into antibody-secreting plasma cells upon interaction with effector CD4+ T-cells. We modeled this interaction using second-order kinetics, where is *T*_BC_ the differentiation rate of B cells into plasma cells.

#### Equation for the activated B cell density (*B*\*(*t*))

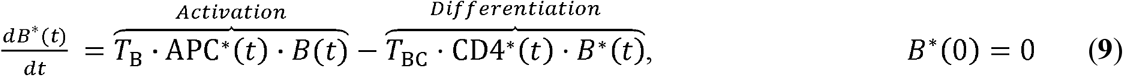

where *T*_BC_ is the differentiation rate of B cells into plasma cells.

#### Equation for the plasma cell density (*P*(*t*))

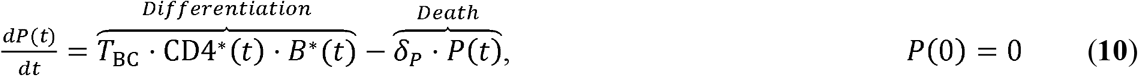

where *δ*_*P*_ is the death rate of plasma cells.

Virus-neutralizing antibodies are secreted by plasma cells, such that their rate of production, characterized by the first-order rate constant *P*_Ab_, is proportional to the plasma cell density. The antibodies secreted into the plasma are then cleared at a rate Cl_Ab_, which is a lumped phenomenological parameter characterizing the various antibody clearance mechanisms.

#### Equation for the neutralizing antibody concentration (Ab(*t*))

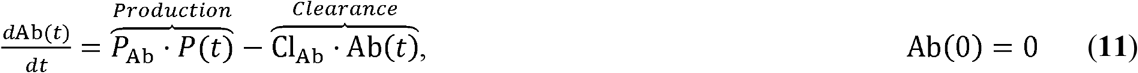

Following vaccination or natural infection, the immune system produces different cytokines to regulate cellular activation and differentiation, as discussed above. In the specific case of SARS-CoV-2, it has been shown that type-I and type-II interferons, and IL-6 are the relevant immune response. For instance, type-I interferons (IFN1(*t*)), secreted by virus-infected cells or immunoregulatory elements [59, 60]. Each cytokine has a unique source and key role in the vaccine-affected cells, lowers the production of new virions by infected cells [52]; type-II interferon (IFN2(*t*)), produced by effector CD4+ and effector CD8+ T-cells, accelerates the differentiation of naive T-cells into their effector form in a positive feedback loop fashion [52]; and IL-6, secreted by effector CD4+ T-cells, effector CD8+ T-cells, and active APCs, tends to exhaust naïve CD4+ and CD8+ T-cell population [58]. The rate of change of cytokine concentration was modeled using a production term and a degradation term, where production and degradation are modeled as first-order processes, with degradation characterized by a common degradation rate constant *δ*_cyt_.

#### Equation for the type-I interferon concentration (IFN1(*t*))

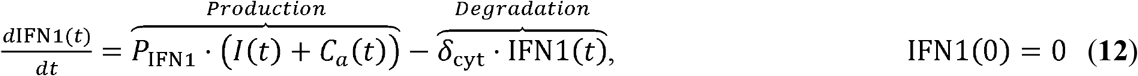

where *P*_INF1_ is the production rate of type-I interferons.

#### Equation for the type-II interferon concentration (IFN2(*t*))

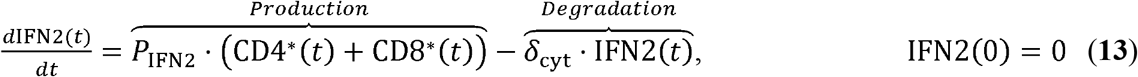

where *P*_INF2_ is the production rate of type-II interferons.

#### Equation for the interleukin-6 concentration (IL6(*t*))

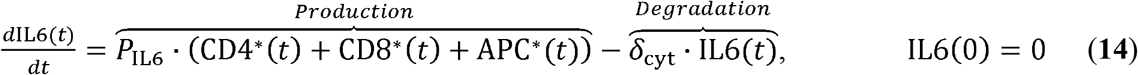

where, *P*_IL6_ is the production rate of IL-6.

The entire immune cascade can be triggered either by a vaccine (as we have already elaborated), or through an infection caused by the SARS-CoV-2 virus. In the latter case, the infection is characterized by the transformation of healthy susceptible cells into infected cells by the virus, followed by production of new viral particles by the infected cells. With the intent to develop a generalized mathematical model capable of simulating immune response to vaccines as well as infections, we incorporate the infection process into our model, with the respiratory tract as a representative site. For this, we used a target cell limited model of acute viral infection [61], as described by the equations below:

#### Equation for the healthy respiratory epithelial cell density (*H*(*t*))

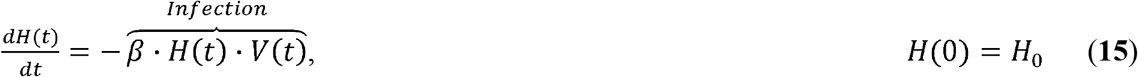

where *β* is the viral infectivity rate, *V*(*t*) is the viral load density in the respiratory tract, and *H*_0_ is the initial density of healthy cells.

#### Equation for the density of infected cells in the respiratory tract epithelium (*I*(*t*))

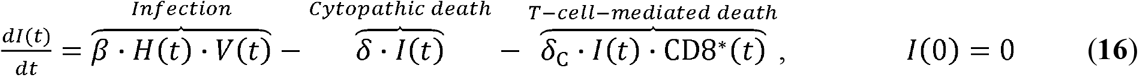

where *δ* represents the cytopathic death rate of infected cells, *δ*_C_ is the death rate of infected cells mediated by effector CD8+ T-cells, and CD8*(*t*) is the density of effector CD8+ T-cells.

#### Equation for the viral load density in the respiratory tract (*V*(*t*))

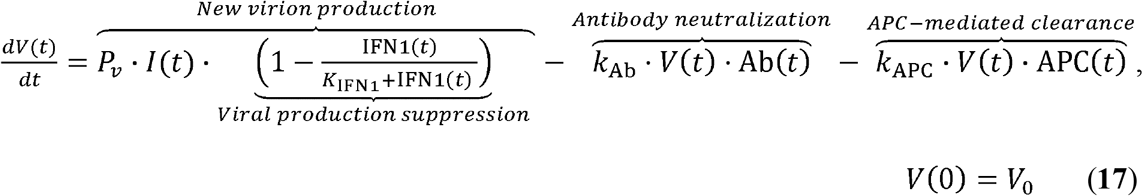

where *P*_*ν*_ represents virion production rate, IFN1(*t*) is the concentration of type-I interferons, *K*_IFN1_ is the Michaelis constant of the virion production suppression factor, *k*_Ab_ is the antibody-mediated neutralization rate of viruses, Ab(*t*) is the antibody concentration in the body, APC(*t*) is the density of naïve APCs in the respiratory tract, *k*_APC_ is the naïve APC-mediated clearance rate of viruses, and *V*_0_ is the initial viral load at the time of infection.

### 2.2 Model calibration and validation

Using the built-in MATLAB function *lsqcurvefit*, non-linear least squares regression was performed to fit the model to literature-derived clinical data to estimate the unknown model parameters (**Table 1**). The datasets used for model calibration included: (i) viral load and immune response kinetics following a SARS-CoV-2 infection [45], (ii) immune response kinetics following vaccination with mRNA vaccines in healthy individuals [46], and (iii) cancer patients undergoing chemotherapy or immunotherapy [47]. Further, to test the predictive ability of our model to accurately reproduce the immune response to mRNA vaccines, we simulated two and three doses of the Pfizer-BioNTech and Moderna vaccines in healthy individuals using the parameters obtained from model calibration for healthy population (**Table 1**), and compared it to published clinical data [46, 62, 63].

Specifically, for calibration of immune response to infection, we digitized average, longitudinal viral load and immune variable data (naïve CD4+ T-cells, naïve CD8+ T-cells, effector CD4+ T-cells, effector CD8+ T-cells, type-I interferons, type-II interferons, IL-6, and neutralizing antibody titer) for moderately infected COVID-19 patients (N = 80 patients) from Lucas et al. [45]. Moderately infected patients were chosen over severely infected ones for the lack of pharmacological intervention in the former, thereby allowing the calibration of purely immune response effects on containing infection. Further, for the calibration of immune response to mRNA-based COVID-19 vaccines in healthy subjects, we extracted average, longitudinal immune response data (neutralizing antibody titer, effector CD4+ T-cells, effector CD8+ T-cells) to two doses of Pfizer-BioNTech COVID-19 mRNA vaccine (N = 31 subjects) from Collier et al. [46]. The two doses were given 21 days apart, with immune response measured 2 to 4 weeks (3 weeks average) after second dose, 6 months after first dose, and 8 months after first dose. From the same study, average immune response data (neutralizing antibody titer) for two doses of Moderna COVID-19 mRNA vaccine (21 days apart) was also extracted for model validation (N = 22 subjects, see below). For calibration of vaccine immune response in cancer patients undergoing antineoplastic treatment, longitudinal antibody titer data (N = 63 patients receiving chemotherapy, N = 16 patients receiving immunotherapy) following two doses of Pfizer-BioNTech COVID-19 mRNA vaccine was extracted from Peeters et al. [47]. The two doses were given 21 days apart and immune response was measured at the time of second dose and 7 and 28 days after second dose. Additional data for model validation (antibody titer kinetics) following two and three doses of Pfizer-BioNTech COVID-19 mRNA vaccine in healthy subjects was obtained from Bayart et al. (N = 158 subjects) [62] and Papazisis et al. (N = 110 subjects) [64], respectively. In Bayart et al., the two doses were given 21 days apart and the immune response was measured 14, 28, 42, 56, 90 and 180 days following the first dose. In Papazisis et al., two doses were given 21 days apart, following which a third dose was given 9 months after the second dose, and immune response was measured 2 weeks after the first dose, 2 weeks after the second dose, 3 months, 6 months, 9 months, and 12 months after the second dose. To account for the uncertainty in parameter estimation during model calibration, model predictions were accompanied by 90% prediction interval obtained through 10,000 simulation runs of the model, where each simulation was obtained for a unique set of parameter values drawn through Latin hypercube sampling (LHS). Note that for LHS, all parameter values were chosen from within a ±10% range of the baseline values estimated during model calibration with healthy subjects.

### 2.3 Vaccine efficacy estimation

In accordance with the literature [65, 66], we used the plasma levels of neutralizing IgG anti-spike antibodies against SARS-CoV-2 as predictors of vaccine efficacy (i.e., correlate of protection against SARS-CoV-2). For this, we characterized an empirical correlation between neutralizing antibody titer (Ab(*t*)) and vaccine efficacy (*V*_eff_(*t*)) based on clinical data from the literature [67]. The following Michaelis-Menten function was thus used:

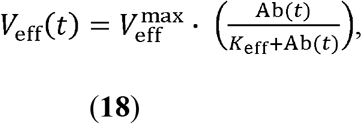

where 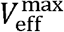 is the maximum possible efficacy of antibody and *K*_eff_ is the Michaelis constant for vaccine efficacy.

As shown in **Figure S1** (solid blue curve), the above function is in excellent agreement with the clinical data, giving an estimate for *K*_eff_ = 18.95 U/mL (i.e., 50% efficacy) and 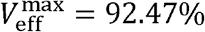. While only 50% efficacy is necessary to obtain approval for clinical use of vaccines [68], our analysis is based on a more stringent threshold to ensure protection in majority of recipients. According to Goldblatt et al. [67], the average plasma antibody titer for various COVID-19 vaccines to be protective against wildtype strain (WT) of SARS-CoV-2 is 154 U/mL, which corresponds to a vaccine efficacy of ∼82.3% on the Michaelis-Menten curve. Therefore, 154 U/mL was used as a threshold to differentiate protected versus non-protected individuals in our analyses. *Note that in this work we assume that individuals with antibody titer above the protection threshold are fully protected, while the ones below are fully at risk of infection* [69].

Of note, for the VOCs, the protective threshold was corrected for by using the binding score (Ab_escape_) of the antibodies obtained from the literature [70]. For this, the previous function was modified to:

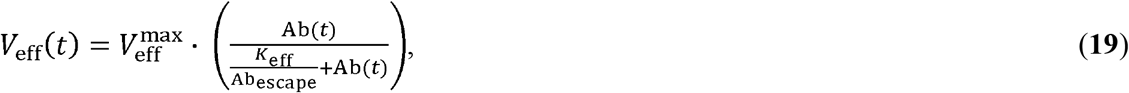

where Ab_escape_ is a dimensionless binding score (Ab_escape_ ∈ [0, 1]) obtained from Greaney et al. that for the VOC [70] that quantifies antibody escape, i.e. the inability of antibodies to neutralize the virus due to inefficient binding. As per Greaney et al., the value of Ab_escape_ for WT is 1 and that for the VOC studied here (i.e., omicron (OM)) is 0.2, indicating that the strength of binding of antibodies to OM is five times lesser than that for WT. As a result, 770 U/mL (i.e., 5 times of 154 U/mL) was the estimated threshold of protection against OM, corresponding to 82.3% vaccine efficacy (based on dotted orange curve in **Figure S1**). Of note, the above calculations assume that the mutations in the RBD- or NTD-domain of SARS-CoV-2 spike protein negatively affect the binding affinity of antibodies [71], which implies that to obtain a similar protection against OM, or other VOCs, a higher antibody titer is necessary.

### 2.4 Sensitivity analysis

To evaluate the relative effect of model parameters on antibody titer following an injection of mRNA-based COVID-19 vaccine, we performed global sensitivity analysis (GSA) and local sensitivity analysis (LSA) with parameters of interest. For this, model parameters were perturbed from their baseline values and the effect of parameter perturbation on model output of interest quantified (i.e., antibody titer). Firstly, to rank order the parameters for their relative importance in determining antibody titer following vaccination, GSA was performed where all model parameters were *simultaneously* perturbed over a uniformly distributed range of ±50% around the baseline parameter values (obtained from model calibration), except parameter *f* that was perturbed between the values of 0.1–1 (left half-Gaussian distribution); area under the antibody concentration kinetics curve (AUC) from zero to 15 days post injection was calculated for each simulation (i.e., for a given combination of parameter values). Note, to comprehensively investigate the vast multiparameter space (22 parameters), yet to minimize the number of simulations, Latin Hypercube Sampling (LHS) was used to obtain 10,000 combinations of parameter values, and 10 such replicates were obtained, based on our previously developed workflows. [72, 73] Multivariate linear regression analysis was then performed on every replicate, and regression coefficients were determined as a quantitative measure of parameter sensitivity index (SI). A distribution of regression coefficients (or SI) was obtained for each parameter, and one-way ANOVA with Tukey’s test was used to rank order the parameters in terms of their sensitivity, such that a higher SI represents a greater influence on model output (i.e., AUC of antibody titer).

Next, to evaluate the correlation between parameter perturbations and change in antibody titer, LSA was performed, where parameters were perturbed individually at 100 linearly spaced levels over a uniformly distributed range of ± 50% around the baseline parameter values (obtained from model calibration). The corresponding change in AUC of antibody titer (from zero to 15 days post injection) with respect to parameter perturbation was calculated with the following formula for SI:

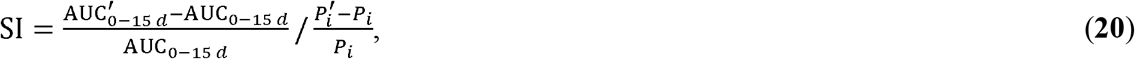

where, AUC_0−15*d*_ is the AUC of antibody titer under baseline conditions from 0-15 days following injection, 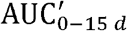 is the AUC of antibody titer under parameter perturbation condition, *P*_*i*_ is the baseline value of parameter *i*, and 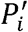 is the perturbed value of parameter *i*.

### 2.5 Virtual patient cohort design

To perform population-scale numerical experiments, two types of patient populations were generated, namely Cohort A and Cohort B, as described below.

#### Cohort A

A virtual cohort of 10,000 individuals was generated using Latin hypercube sampling (LHS) [74-76] from twelve parameter distributions (**Figure S2**), such that each individual of the cohort varied in terms of their immune health status defined by *f*, underlying biology (characterized by the higher-ranking parameters of GSA, i.e., top 10 parameters), and vaccination schedule. The chosen range for the parameter values was such that the *f* parameter varied between 0.5 and 1 (left half-Gaussian distribution with mode equal to 1 and standard deviation equal to 15% of the mode, **Figure S2c**), while the other biological parameters were normally distributed with a mean equal to the baseline parameter value and one standard deviation equal to 5% of the mean value (**Figure S2d-l**); the dosing schedules varied between two weeks and eight weeks for the second dose (continuous uniform distribution, **Figure S2a**), and between five months and nine months for the first booster dose (i.e., third dose; continuous uniform distribution, **Figure S2b**). Note that to reflect the distribution of immune health status and biological variability in a population realistically, i.e., *majority* of the population comprising healthy individuals, Gaussian distributions with limited variance were chosen over unform distributions for LHS.

#### Cohort B

Three virtual sub-cohorts of 10,000 individuals each to represent healthy, mildly immunocompromised, and highly immunocompromised individuals were generated through LHS. The range of *f* values used to represent immune health status was *f* = 0.9 to 1 for healthy (continuous uniform distribution), *f* = 0.7 to 0.9 for mildly immunocompromised (continuous uniform distribution), and *f* = 0.5 to 0.7 for highly immunocompromised individuals (continuous uniform distribution). Also, biological variability was included in each sub-cohort by LHS of the relevant biological parameters (identified through GSA, same as used for cohort A), assumed to be normally distributed with a mean equal to the baseline parameter value and one standard deviation equal to 5% of the mean value. For each sub-cohort, we tested 100 dosing schedules ranging from two to eight weeks (after the first dose) for the second dose (continuous uniform distribution), 0.5 to nine months (after the second dose) for the first booster (i.e., third dose) (continuous uniform distribution), and one to nine months (after the first booster) for the second booster (i.e., fourth dose; continuous uniform distribution) for their effect on continuity of protection against OM (i.e., vulnerability to breakthrough infection due to mutants).

### 2.6 Vulnerability kinetics and vaccine dosing schedule optimization

To study the temporal evolution and quantify the vulnerability to breakthrough infections at the population scale, we calculated a vulnerability kinetics curve in our numerical experiments (as shown in **Figure 6c**). From the vaccine efficacy calculation (based on **Figure S1**), on a given day, the fraction of simulated individuals below the protection threshold for OM or WT (i.e., <82.3% efficacy) was calculated to obtain the population fraction that is at a high risk of infection. Performing this calculation daily for the entire simulation period gave us the curve shown in **Figure 6c**, referred to as the vulnerability kinetics curve. Subsequently, we calculated the area under the curve (AUC) as a measure of total vulnerability to breakthrough infections, which was then used as a metric for optimizing dosing schedules to impart prolonged protection against OM, as discussed below. Note that we do not use epidemiological principles to model the spread of infection or risk of exposure amongst the individuals of virtual cohorts, given that the goal of our work is to only evaluate the effect of dosing schedule optimization in maintaining protection in already exposed individuals.

To optimize the timing of the second dose, immune response kinetics for each virtual individual (Cohort B) was simulated for up to 150 days after the first dose (given on day 0). From the corresponding antibody concentration kinetics, the vaccine efficacy kinetics for OM were computed using **Eq. 19**. Subsequently, we estimated the vulnerability to breakthrough infections over time. From the vulnerability kinetics plot, the area under the curve (AUC_0-150d_) was calculated using the trapezoidal method. After calculating the AUC_0-150d_ for 100 dosing schedules (ranging from two to eight weeks for the three cohorts), we identified the schedules that led to a minimum in the three sub-populations, which translates to a minimized vulnerability to breakthrough infections resulting from OM.

Next, using the optima found in the previous step, we repeated the process to identify the optimal timing for the first booster (third dose) in the three cohorts. In this case, the total simulated time was 500 days. Thus, the AUC_0-500 d_ was calculated from the breakthrough infection vulnerability kinetics plots to identify the minima. Finally, using the optimal dosing schedules for the second dose and first booster (third dose), we estimated the optimal timing for the second booster (fourth dose) using the same process as described before. In this case, the total simulation time was 700 days.

## 3. Results and Discussion

### 3.1 Model calibration

The focus of this work was to mechanistically model the individual-scale immune response to COVID-19 vaccines and apply it to optimize vaccine dosing schedules to maximize protection against SARS-CoV-2 and thus minimize breakthrough infections in the population. For this purpose, we began by fitting the model to immune response kinetics of SARS-CoV-2 infection [45], which allowed us to estimate several unknown model parameters relevant to key immune response variables that were otherwise difficult to compute from vaccination data alone (**Table 1**). This enabled the reliable simulation of immune response kinetics following infection. As shown in **Figure 2a**, the numerical solutions of the model are in agreement with the clinical data for viral load and immune response kinetics following SARS-CoV-2 infection [45]. This is also indicated by the strong Pearson correlation between the observations and the model fits (**Figure S3a**; *R* > 0.99). The computed kinetics of viral load in the respiratory tract predict an incubation period of eight to nine days, which is in accordance with values established in the literature [77]. Moreover, the simulations closely approximated the kinetics for eight additional cellular and molecular immune response variables, including naïve and effector lymphocytes, antibodies, interferons, and interleukins. This suggests that the model predictions are within physiological limits and thus the estimated parameter values are reliable. The results also showed that the viral load peaks around day 10, reaching a level of ∼10^7^GE·mL^−1^ while adaptive immunity variables (lymphocytes, neutralizing antibodies) peaked at around day 15, which led to clearing of the infection within five weeks without any pharmacological intervention.

**Figure 2.**
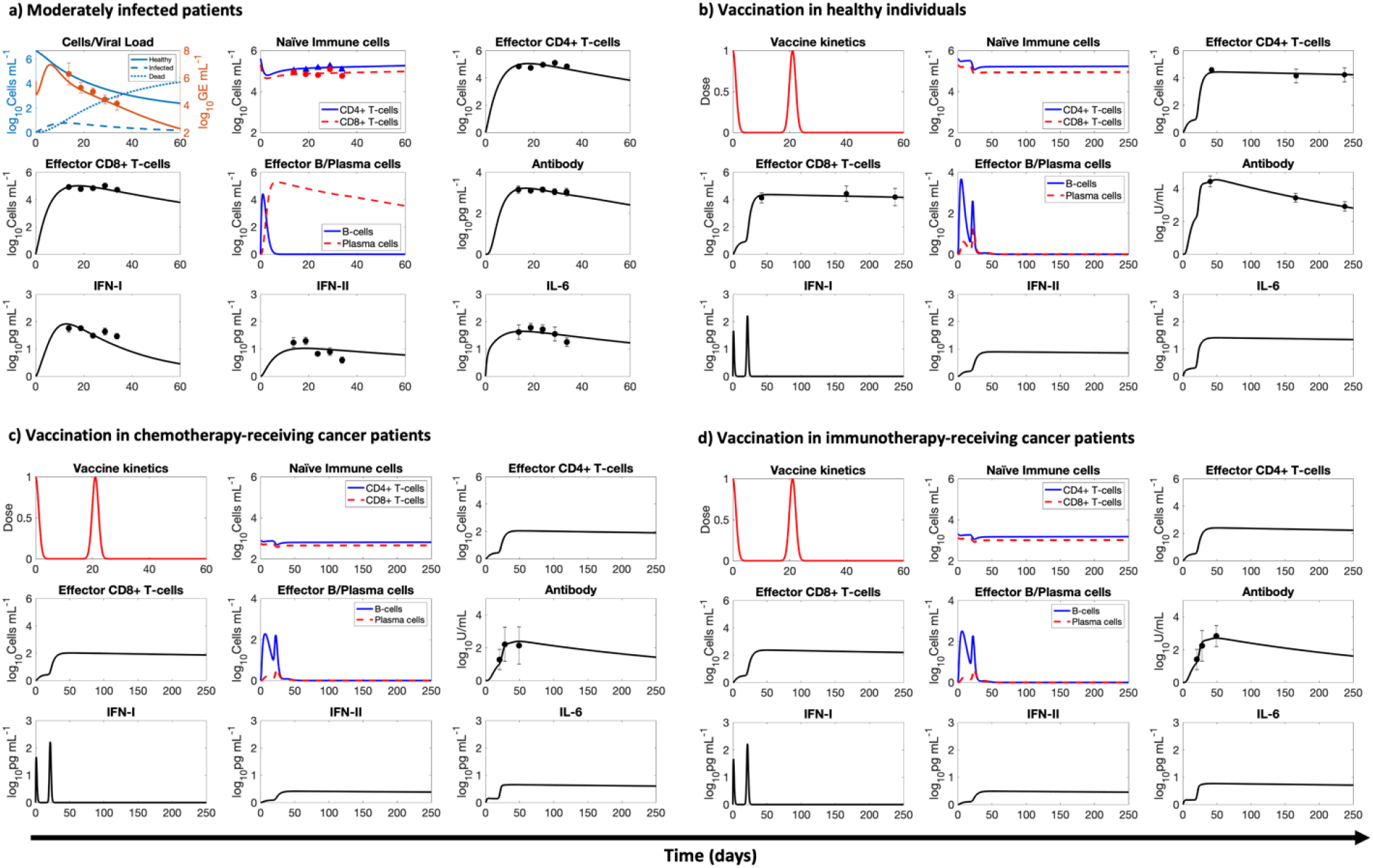
Model calibration. Model calibration with literature-derived clinical data of immune system response kinetics during **a**) SARS-CoV-2 infection in moderately infected subjects, vaccination in **b**) healthy individuals, **c**) cancer patients receiving chemotherapy, and **d**) cancer patients receiving immunotherapy. For consistency, all immunization data was based on two simulations; markers with error bars represent mean ± standard deviation values. doses of the Pfizer-BioNTech COVID-19 mRNA vaccine. Solid or dashed lines indicate model simulations; markers with error bars represent mean ± standard deviation values.

Subsequently, as shown in **Figure 2b**, we calibrated the model with the clinical data obtained from healthy individuals vaccinated with mRNA vaccines (specifically Pfizer-BioNTech) [46]. For this purpose, a double dose of the vaccine was simulated in accordance with the schedule used for the individuals in the study [46]. A Gaussian function described the kinetics of antigen load following injections on days 0 and 28 (**Eq. 1**). The solutions for the various immune response variables were computed over a period of eight months and fitted to the available clinical data for effector T-cells (CD4+ and CD8+) and neutralizing antibodies. Our results showed a high degree of correlation between the model fits and clinical measurements (**Figure S3b**; *R* > 0.96). To ensure that the model can reproduce immune responses elicited by the vaccines over long time periods, some of the parameters were refitted (**Table 1**). Since during the previous calibration, the characteristic time of simulation is a few weeks unlike the current simulation where the simulated time is a few months, we recalibrated some parameters to ensure long term accuracy of the simulation. Also, to adjust the model for it to be able to capture any fundamental differences between response to infection and vaccines, we performed the recalibration. In addition, some parameters required recalibration because of the variation in units of measurement between experiments. An important observation is the gradually waning levels of neutralizing antibodies and effector lymphocytes, which suggests that protection conferred by mRNA vaccines is temporal, warranting the use of boosters.

To accurately represent the vaccine-induced immune response in immunocompromised individuals, we also calibrated the model with clinical data obtained from vaccinated cancer patients undergoing chemotherapy or immunotherapy (**Figure 2c,d**) [47]. In both cases, we assumed that due to the underlying pathophysiology and associated treatment, the levels of some immune system parameters were only a fraction (0 < *f* < 1) of their values in healthy individuals (*f* = 1). Therefore, keeping all other model parameters from the previous two fits as constants, we fitted the model to two datasets [47] to estimate the parameter *f*, which resulted in a value of *f* = 0.517for chemotherapy-treated patients and *f* = 0.588 for immunotherapy-treated cancer patients. The model fits were also in good agreement with clinical data (**Figure S3c,d**; *R* > 0.96).

### 3.2 Model validation

To test the ability of our model to accurately reproduce the immune response to mRNA vaccines, we simulated two and three doses of the Pfizer-BioNTech and Moderna COVID-19 vaccines in healthy individuals (data not used for calibration). As shown in **Figures 3** and **S4** (*R* = 0.92), the computed neutralizing antibody (IgG) kinetics closely resembles the literature-derived clinical data following two doses of the Pfizer-BioNTech COVID-19 vaccine [62], two doses of the Moderna COVID-19 vaccine [46], and three doses of the Pfizer-BioNTech COVID-19 vaccine [78]. The dosing schedules were obtained from the respective clinical studies, and the parameter values were based on the values calibrated for healthy individuals in the previous section (**Table 1**). The ability of the model to accurately predict the response to the third dose, despite not using the third dose data during model calibration, highlights the biological and physiological robustness of our mechanistic model. Having established the validity of our model to reliably reproduce neutralizing antibody kinetics with various mRNA vaccines and dosing schedules, we proceeded to perform numerical experiments to explore the heterogeneity in immune responses and optimize dosing schedules to minimize breakthrough infections.

**Figure 3.**
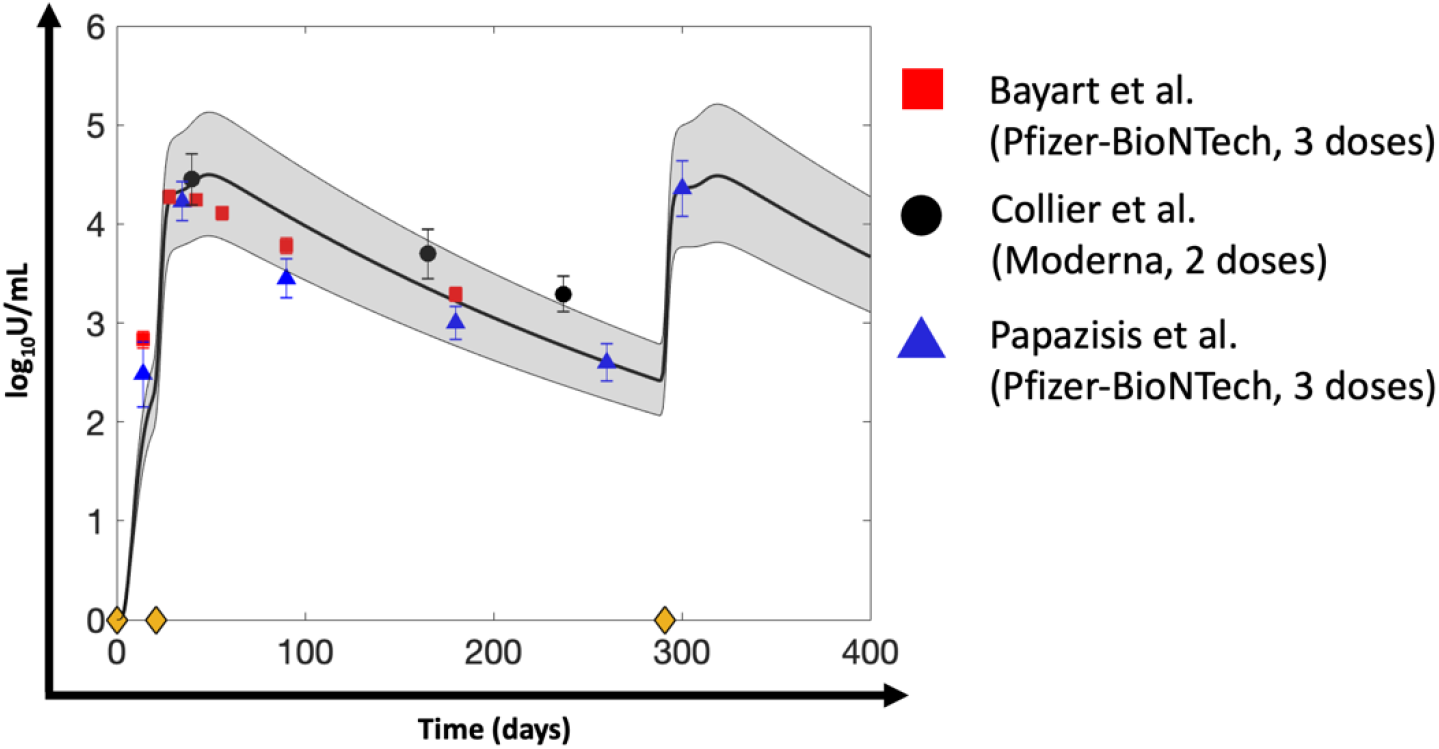
Model validation. Validation of the mathematical model with antibody kinetics data derived from the literature for healthy individuals vaccinated with two doses of Pfizer-BioNTech COVID-19 mRNA vaccine (red squares), two doses of Moderna COVID-19 mRNA vaccine (black circles), and three doses of Pfizer-BioNTech COVID-19 mRNA vaccine (blue triangles). Solid line indicates model predictions, grey bands represent 90% prediction intervals, and markers with error bars represent mean ± standard deviation values of clinical data. Yellow diamonds on the x-axis denote timing of injection (i.e., first dose given on day 0, second dose given on day 21 after first dose, and third dose given 9 months after second dose).

### 3.3 Sensitivity analysis

To study the relative significance of model parameters in governing humoral response to mRNA-based COVID-19 vaccines, quantified as area under the curve of neutralizing antibody titer kinetics following a unit dose of the vaccine, global sensitivity analysis (GSA) and local sensitivity analysis (LSA) were performed with 22 model parameters that characterize the key immunological interactions, processes, and immune states considered to be important for vaccine-induced immune response generation. As shown in **Figure 4**, the immunosuppression factor *f* stands out as the most relevant parameter in determining antibody response to vaccines, which highlights the importance of immune health status (i.e., immune cell sufficiency) in governing vaccine-induced protection. Further, as shown in the inset of **Figure 4**, LSA reveals a positive monotonic correlation between change in *f* and sensitivity index (SI) for antibody titer, which signifies better antibody response in immunologically competent subjects, and thus warrants the need for additional doses or optimized dosing frequency in immunocompromised patients [8]. Following this, biological parameters that characterize antibody production *P*_Ab_, death of antibody secreting plasma cells *δ*_*P*_, and antigen-induced activation of naïve APCs, *T*_APC_ were observed to also influence antibody response strongly (**Figure 4**), thereby indicating the relevance of antigen presentation, plasma cell population, and antibody secretion from plasma cells in humoral immunity development. While the relationship between change in antibody titer and perturbations of parameters *P*_Ab_ or *T*_APC_ is monotonically increasing within the studied parameter range, that between antibody titer and *δ*_*P*_ is monotonically decreasing (**Figure 4, inset**). Further, additional parameters belonging to CD4+ T-cell activation *T*_CD4_, growth of B cells *γ*_*B*_ death of activated APCs *δ*_APC_, differentiation of B cells into antibody secreting plasma cells *T*_BC_, potency of the injected antigen to activate naïve APCs, v, and activation of naïve B cells, are also observed to have a moderately significant effect on antibody titer (**Figure 4**). This further confirms the relevance of previously identified processes, in addition to the levels of activated CD4+ T-cells and B cells in governing antibody response. While the parameters identified in this analysis do not contribute directly to immune health status in our model, except *f*, their significance in antibody response warrants their inclusion in virtual patient cohort generation to study the effect of the underlying population-scale biological variability on vaccine-induced protection.

**Figure 4.**
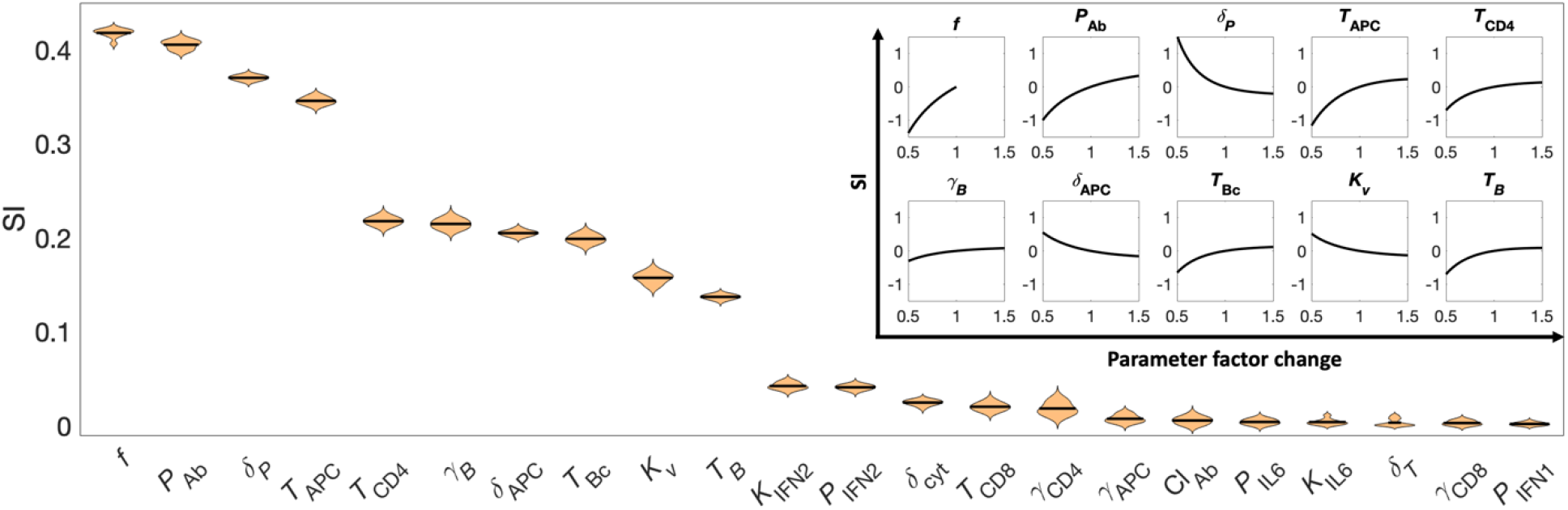
Parameter sensitivity analyses. Violin plot showing parameters ranked in descending order (from left to right) for their sensitivity, as indicated by the sensitivity index (SI) obtained through global sensitivity analysis (GSA). Correlation of parameter perturbation and its effect on antibody titer (quantified as SI) obtained through local sensitivity analysis (LSA) for the top ten ranking parameters of GSA is shown in the inset. Note that parameters were perturbed in the range of ± 50% around their baseline values for both GSA and LSA.

### 3.4 Heterogeneity in immune response to vaccines at the individual and population scale

To study the influence of (i) vaccine dosing schedules and (ii) the immune status of an individual on neutralizing antibody levels and vaccine efficacy, we simulated immune responses under different dosing schedules in representative healthy and immunocompromised subjects. Based on the dosing schedules used across various countries, we considered three vaccination regimens: *rapid, intermediate*, and *delayed*. In all cases the first dose was given on day 0; (i) Rapid: second dose is given two weeks after the first dose, and the first booster (third dose) is given five months after the second dose; (ii) Intermediate: second dose is given four weeks after the first dose, and the first booster is given seven months after the second dose; (iii) Delayed: second dose is given eight weeks after the first dose, and the first booster (third dose) is given nine months after the second dose.

Here, the immune health status was defined by the non-dimensional, empirical parameter *f*, such that healthy individuals have *f* = 1, mildly immunocompromised subjects have *f* = 0.75, and highly immunocompromised individuals have *f* = 0.55. As previously discussed, *f* = 0.55 corresponds to cancer patients undergoing chemotherapy or immunotherapy, whereas *f* = 0.75 simulates individuals with underlying conditions that may also affect the immune system, but usually to a lesser degree, (e.g., autoimmune diseases). As per evidence in the literature, plasma antibody titer is a correlate of protection against infection [65, 66]. Therefore, we used the computed neutralizing antibody levels as predictors of vaccine efficacy (i.e., protection against SARS-CoV-2; see section 2.3 in **Methods**).

We used the model to predict the humoral response to mRNA vaccines following three dosing schedules in representative healthy or immunocompromised individuals for a 600-day period. As shown in **Figure 5a,d,g** (upper subplot in each panel), irrespective of the dosing schedule, the antibody levels remain above the protection threshold for both OM (770 U/mL) and WT strain (154 U/mL) for a much longer duration in healthy individuals (as indicated by the shaded grey area and quantified as the *T*_safe_ value = 383–443 days) than in mildly immunocompromised subjects (**Figure 5b,e,h**; *T*_safe_ = 162–228 days). In contrast, in cancer patients undergoing antineoplastic treatment (i.e., highly immunocompromised subjects), *T*_safe_ was zero days across all dosing schedules (**Figure 5c,f,i**). This suggests that highly immunocompromised subjects are vulnerable to infection with OM throughout the 600-day simulation period, however protection against WT is intermittently present depending upon the dosing schedule. Of note, within both the healthy and mildly immunocompromised subjects, the intermediate dosing schedule leads to higher *T*_safe_ values (443 days if healthy, 228 days if mildly immunocompromised; **Figure 5d,e**) than the rapid dosing schedule (383 days if healthy, 162 days if mildly immunocompromised; **Figure 5a,b**) or the delayed dosing schedule (396 days if healthy, 182 days if mildly immunocompromised; **Figure 5g,h**). Nonetheless, the protection window in these cases is not continuous for the chosen dosing schedules, and an intermediate ‘gap’ is observed between the second dose and first booster (third dose) that highlights the period when antibody levels temporarily fall below the protective threshold for OM and/or WT. The duration of this gap varies according to immune health status and dosing schedule.

**Figure 5.**
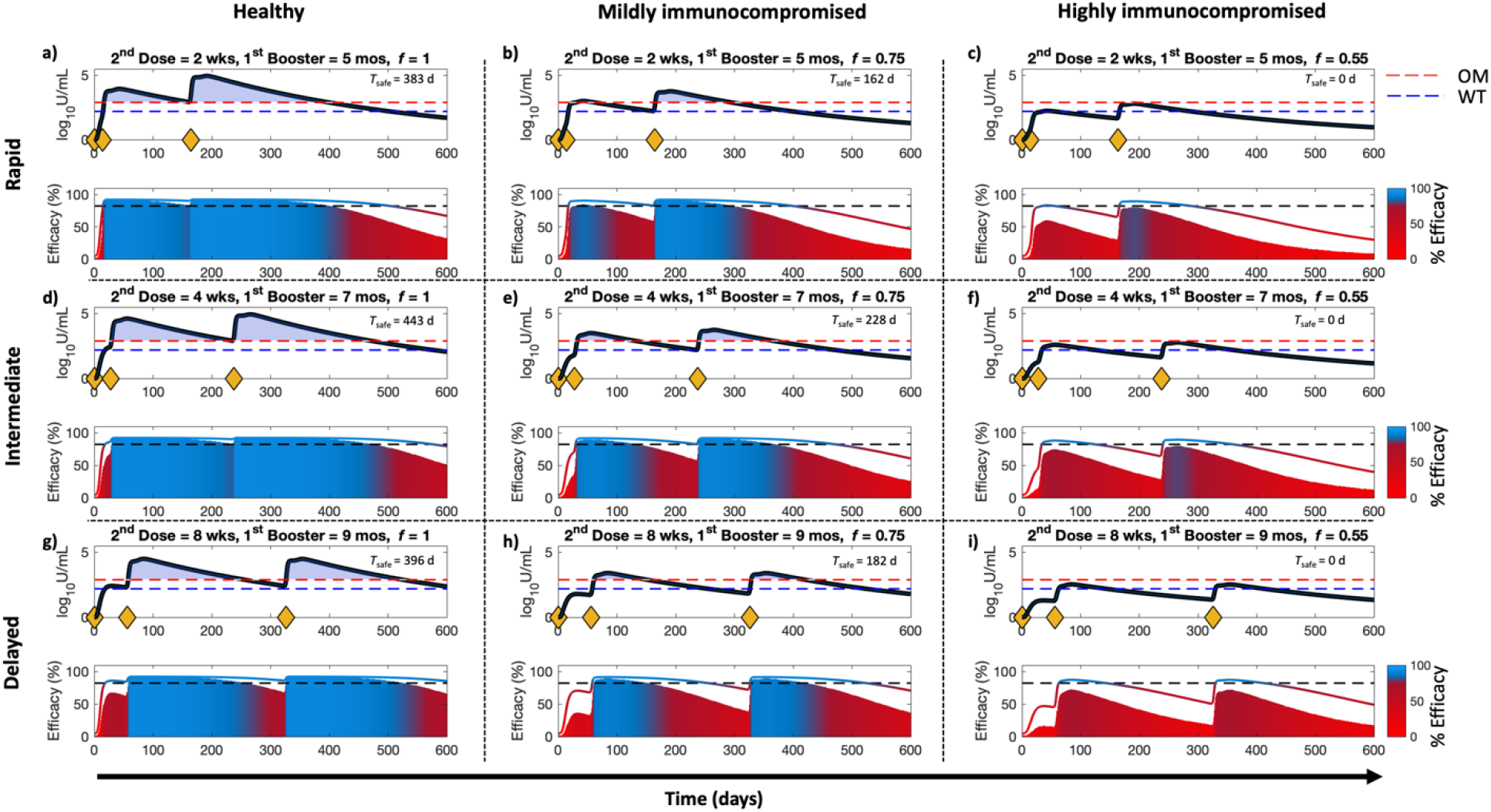
Effect of vaccine dosing schedule and immune health status on antibody levels and vaccine efficacy. Simulations in representative (**a,d,g**) healthy and (**b,c,e,f,h,i**) immunocompromised individuals show antibody levels and vaccine efficacy against wildtype strain (WT) and Omicron variant (OM) of SARS-CoV-2 following (**a,b,c**) rapid, (**d,e,f**) intermediate, and (**g,h,i**) delayed vaccine dosing schedules. Yellow diamonds on the x-axes, in the upper sub-panel, indicate injection timepoints. In each upper sub-panel, the black solid line represents antibody levels, with the dashed blue and red lines indicating protective threshold against WT and OM, respectively. The lower sub-panel shows vaccine efficacy (colored solid line for WT and shaded area for OM), with the dashed black line indicating the 82.3% threshold of protection. Note: The value *T*_safe_ indicated in every upper sub-panel represents the number of days when antibody levels are above the protective threshold for both WT and OM.

The corresponding vaccine efficacy kinetics are shown in the lower subplots in **Figure 5**. The shaded area represents the vaccine efficacy against OM, and the solid-colored line indicates the vaccine efficacy against WT. The continuous color mapping assigns blue to efficacies above the protection threshold (>82.3%) and red to efficacies equal to or below the protection threshold (≤ 82.3%). As visible from the bluish region of the shaded area, for any given dosing schedule, protection threshold healthy individuals have greater vaccine efficacy against OM than immunocompromised individuals. In highly immunocompromised subjects the shaded area always remained below the protective threshold (82.3%), indicating a high risk of becoming infected with OM (**Figure 5c,f,i**). As expected, due to limited *antibody escape* [79], the vaccine efficacy against WT is greater than that against OM in all individuals under all dosing schedules (as indicated by the colored solid line). Further, in healthy individuals, the three dosing schedules produced antibody titers above the WT protection threshold for the majority of the simulation period (**Figure 5a,d,g**). In mildly immunocompromised individuals, protection against WT does not persist continuously (**Figure 5b,e,h**). For example, in the delayed dosing schedule shown in **Figure 5h**, the period between day 236 and day 330 (∼three months) indicates a vaccine efficacy of less than 82.3%. In highly immunocompromised cases, the three dosing schedules provide limited protection against WT, with prolonged periods of lapse in immunity. Though we only considered representative individuals, these observations collectively highlight the importance of optimizing the dosing schedule based on the immune health status of a sub-population to achieve continuous, long-term protection against both WT and other VOCs (e.g., OM).

To evaluate the effects of dosing schedules and immune health status on the variability in immune response to mRNA vaccines at the population level, we simulated the vaccination of a virtual population of 10,000 individuals with three doses (**Cohort A**; see section 2.5 in **Methods** for details of dosing schedule) and assessed the corresponding vulnerability to breakthrough infections. Note that the dosing schedule for each simulated individual was obtained randomly from a continuous time interval (red and blue brackets on x-axis of **Figure 6**) to replicate the real-world heterogeneity in dosing time intervals. As shown in **Figure 6a**, the average antibody kinetics across the 10,000 individuals remained above the protective threshold for OM and WT. However, for a significant fraction of the population, antibody levels remained below the OM threshold for a prolonged period (∼five months). This is evident from the shaded area representing 90% prediction interval. Further, translating the antibody levels to vaccine efficacy using **Eq. 19**, we observed that for a significant fraction of the 10,000 individuals, vaccine efficacy against OM fell below the 82.3% protection threshold (see **Figure 6b**, orange shaded area). Subsequently, we quantified the fraction of the virtual population that presented a vaccine efficacy below the protective threshold for OM and WT (**Figure 6c**, see section 2.6 in **Methods)**. This population fraction can alternatively be interpreted as the fraction of vaccinated individuals in a population that is vulnerable to breakthrough infections, i.e., becoming infected despite being vaccinated. As observed in **Figure 6c**, this fraction increases to about 0.5 (or ∼50% of the population) for OM in vaccinated individuals (two doses), and then declines rapidly following administration of the first booster (third dose). However, due to waning antibody levels, which translate into declining efficacy, the vulnerable fraction begins to increase again and becomes 1 (i.e., 100% of population) in about six months after the booster window. In contrast, for WT, the vulnerable fraction of the population peaks at about 0.1 (or ∼10% of the population) in vaccinated individuals (two doses) and then decreases again after administration of the first booster (third dose), suggesting effective protection against WT in vaccinated individuals for up to ∼1.5 years, irrespective of the dosing schedule or immune health status. Of note, in the population-scale simulation, immune health status is non-uniformly distributed across the population, as defined by the left half-Gaussian distribution (**Figure S2c**); this indicates that a major proportion of the population is healthy. It is worth mentioning that the sharp rise in **Figure 6c** of the population fraction several months after the first booster warrants the administration of a second booster to curb the vulnerability to VOCs and WT. Given that the proposed dosing schedules do not warrant continuous protection against VOCs and/or WT, it is imperative to optimize the schedules to achieve long-term protection in the population without lapses.

**Figure 6.**
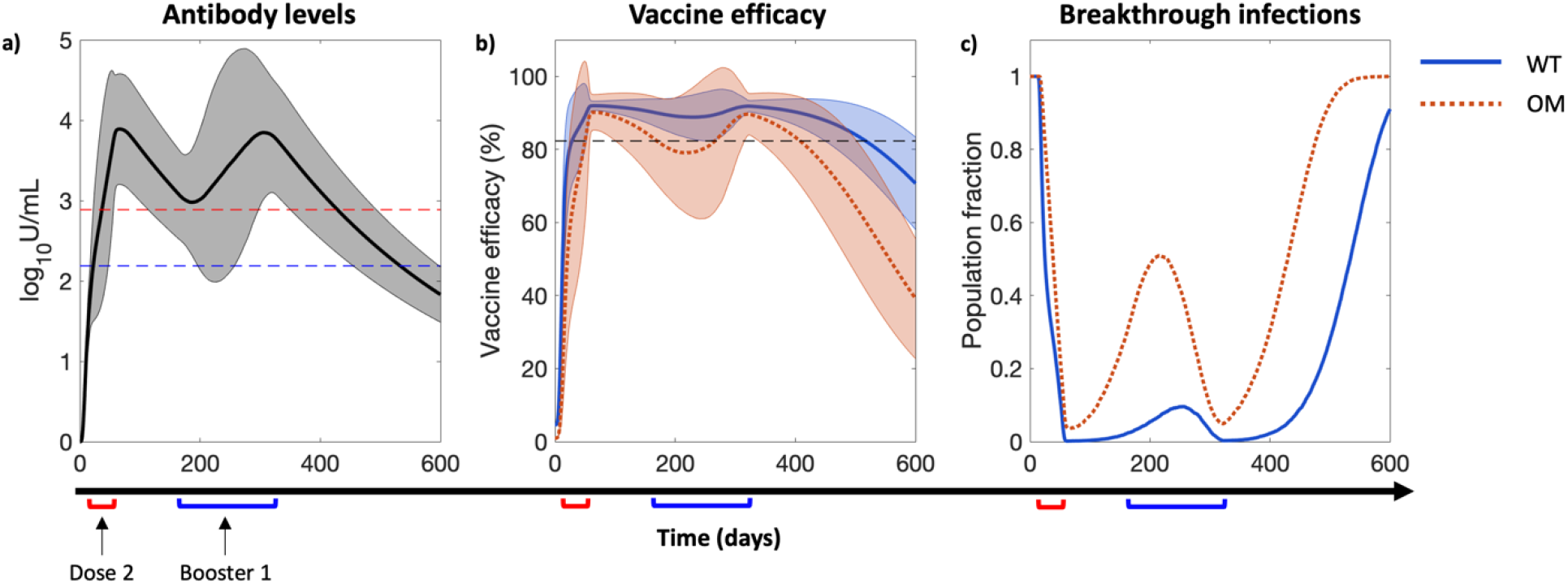
Effect of heterogeneity in vaccine dosing schedules and immune health status on breakthrough infections at the population scale. **a**) Average antibody levels in plasma, **b**) corresponding vaccine or antibody efficacy, and **c**) population fraction vulnerable to breakthrough infections due to wildtype strain (WT, solid blue line) and Omicron variant (OM, dotted orange line) of SARS-CoV-2 over time. Solid and dotted lines in **a,b**) represent average behavior of 10,000 simulated individuals and shaded bands indicate 90% prediction interval. Note that the first dose was administered on day 0 to each simulated individual, the second dose was administered between day 14 and day 56, and the third dose (i.e., first booster) was administered between day 150 and day 270. Red and blue brackets on x-axis denote timing windows with respect to day 0 for second dose and third dose, respectively, used to design unique vaccine schedules in model simulations. Immune health status (*f*) of the simulated population varied between 0.5 to 1.

### 3.5 Vaccine dosing schedule optimization

Following the previous numerical experiments, we intended to identify optimal vaccine dosing schedules to achieve continuous protection against OM (as a representative example) for prolonged periods. We generated three virtual cohorts of 10,000 individuals (**Cohort B**) each to represent healthy, mildly immunocompromised, and highly immunocompromised individuals, and implemented several dosing schedules to identify optimal times for the second dose, the third dose (first booster), and the fourth dose (second booster) in each sub-cohort (see section 2.6 in **Methods**).

As shown in **Figure 7**, the AUC of vulnerability kinetics curves follows a non-linear relationship with respect to dosing schedules, and a minimum is visible for each dose and population sub-type (highlighted by a red circle). As shown in **Figure 7a,d,g**, as the immune status changes from healthy to highly immunocompromised, the position of the minima on the x-axis shows a right shift, such that the optimal time for the second dose in healthy, mildly immunocompromised, and highly immunocompromised individuals is 18 days, 25 days, and 30 days after the first dose, respectively. In contrast, as shown in **Figure 7b,e,h**, the minima for the first booster shows a left shift on the x-axis from healthy to highly immunocompromised individuals, such that the optimal time for first booster is 164 days (∼5.5 months), 115 days (∼4 months), and 36 days (1.2 months) after the second dose for healthy, mildly immunocompromised, and highly immunocompromised individuals, respectively. Similarly, as shown in **Figure 7c,f,i**, the minima for the 2^nd^ booster shows a left shift from healthy to highly immunocompromised individuals, such that the optimal schedule for the second booster is 223 days (∼7.5 months), 195 days (∼6.5 months), and 126 days (∼4 months) after the first booster for healthy, mildly immunocompromised, and highly immunocompromised individuals, respectively.

**Figure 7.**
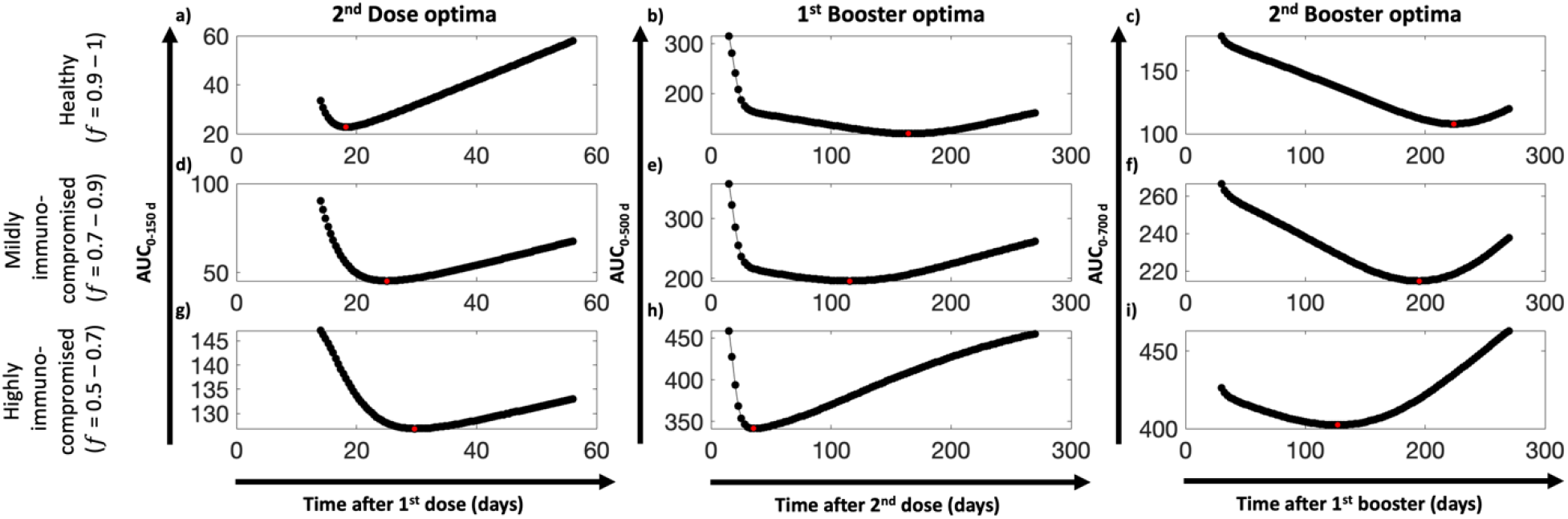
COVID-19 vaccine dosing schedule optimization. Area under the curve (AUC) of breakthrough infection vulnerability kinetics curve obtained from simulation of 10,000 individuals from different population sub-types under unique dosing schedules and immune health status. Estimated AUC versus dosing schedules for **a,d,g**) dose 2, **b,e,h**) booster 1 (i.e., dose 3), and **c,f,i**) booster 2 (i.e., dose 4) for **a,b,c**) healthy, **d,e,f**) mildly immunocompromised, and **g,h,i**) highly immunocompromised individuals, obtained through model simulations. Each black dot represents one AUC value. Red dot in each plot represents the corresponding minima for each dose and population sub-type.

It is intuitive to expect inter-vaccination periods to be longer for healthy individuals than for immunocompromised patients. This is evidenced by data presented in **Figure 5**, where the antibody titer stays above the OM protection threshold for a longer period in healthy individuals than in their immunocompromised counterparts, thereby allowing the possibility to delay subsequent doses. Although this is true for the first and the second boosters (**Figure 7b** vs. **Figures 7e,h**; **Figure 7c** vs. **Figures 7f,i**), the trend is reversed for the second dose (**Figure 7a** vs. **Figures 7d,g**), where healthy individuals seem to require the second dose sooner than immunocompromised individuals to ensure continuity of protection against OM. This observation can be explained in light of a key mechanistic assumption of our model. Recall that the immune health status parameter *f* scales the homeostasis level of naïve immune cells (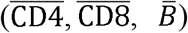). In immunocompromised individuals, *f* ranges from 0.5 to 0.9; therefore, the homeostasis level of naïve immune cells is less than that in healthy individuals (**Figures 2b,c,d**). As a result, when the second dose is given too soon after the first dose in immunocompromised individuals, due to reduced levels of CD4+ T-cells and therefore slower activation of B cells, the production of neutralizing antibodies from plasma cells may be thwarted, thereby rendering an individual vulnerable to infection. Therefore, permitting the CD4+ T-cell and B cell population to regenerate after the first dose will allow antibody titers to rise to levels associated with adequate protection. Of note, since healthy individuals produce or activate immune cells more quickly (given *f* = 1), they are ready to receive a second dose sooner than immunocompromised individuals. However, in the case of healthy individuals, as shown in **Figure 7a**, the AUC_0-150 d_ values are smaller than those of immunocompromised patients (**Figure 7d,g**) for up to ∼six weeks of delay after the first dose. A six-week delay after the first dose predisposes ∼30% (obtained from the ratio of AUC_0-150 d_ value at six weeks (i.e., ∼45) to maximum possible value of AUC_0-150 d_ (i.e., 150)) of the healthy population to a breakthrough infection over 150 days under no public health restrictions. This indicates that although an optimal waiting period for healthy individuals is two and a half weeks after the first dose (which predisposes only ∼15% healthy population over 150 days), if required due to logistic constraints, waiting longer (up to six weeks) will still allow the healthy individuals to be more protected than immunocompromised individuals.

### 3.6 Testing model-predicted optimal dosing schedules

Finally, to demonstrate the impact of the previously identified optimal dosing schedules (for the second dose and the two boosters) in reducing vulnerability to breakthrough infections, we simulated a vaccination regimen with four doses in 10,000 virtual individuals per group, belonging to the three cohorts of interest (healthy, mildly immunocompromised, and highly immunocompromised; see **Cohort B** in section 2.5 in **Methods**), and measured the vaccine efficacy and corresponding level of vulnerability to infection over a period of two years. Note that to simulate a more realistic test scenario and add variability to the optimal dosing schedules identified previously (**Figure 7**), we sampled the dosing schedules from within a ± 10 % uniform distribution around the optimal values. As shown in **Figures 8a,c,e**, the average vaccine efficacy for WT and OM is above the protection threshold in all sub-populations for an extended period of time, the duration of which is dependent on the viral strain and population sub-type. Therefore, the corresponding vulnerability to breakthrough infections for OM and WT remains at almost zero for most of the two-year period in healthy individuals and shows only two intermittent windows of ∼two months each where the vulnerability is as high as ∼0.065 (**Figure 8b**). In mildly immunocompromised individuals, the optimized protocol exhibits similar results, although the vulnerability to infection after fourth dose begins to rise sooner in comparison to the healthy population (**Figure 8d**). Furthermore, as shown in **Figure 8f**, in the highly immunocompromised cohort, the same trend continues; although complete protection against OM and WT is observed for a shorter duration, the results are nonetheless notably more promising compared to the observed findings in **Figures 5c,f,i**, where vaccine efficacy remained below the OM protection threshold throughout the 600-day window under conventional dosing schedules.

**Figure 8.**
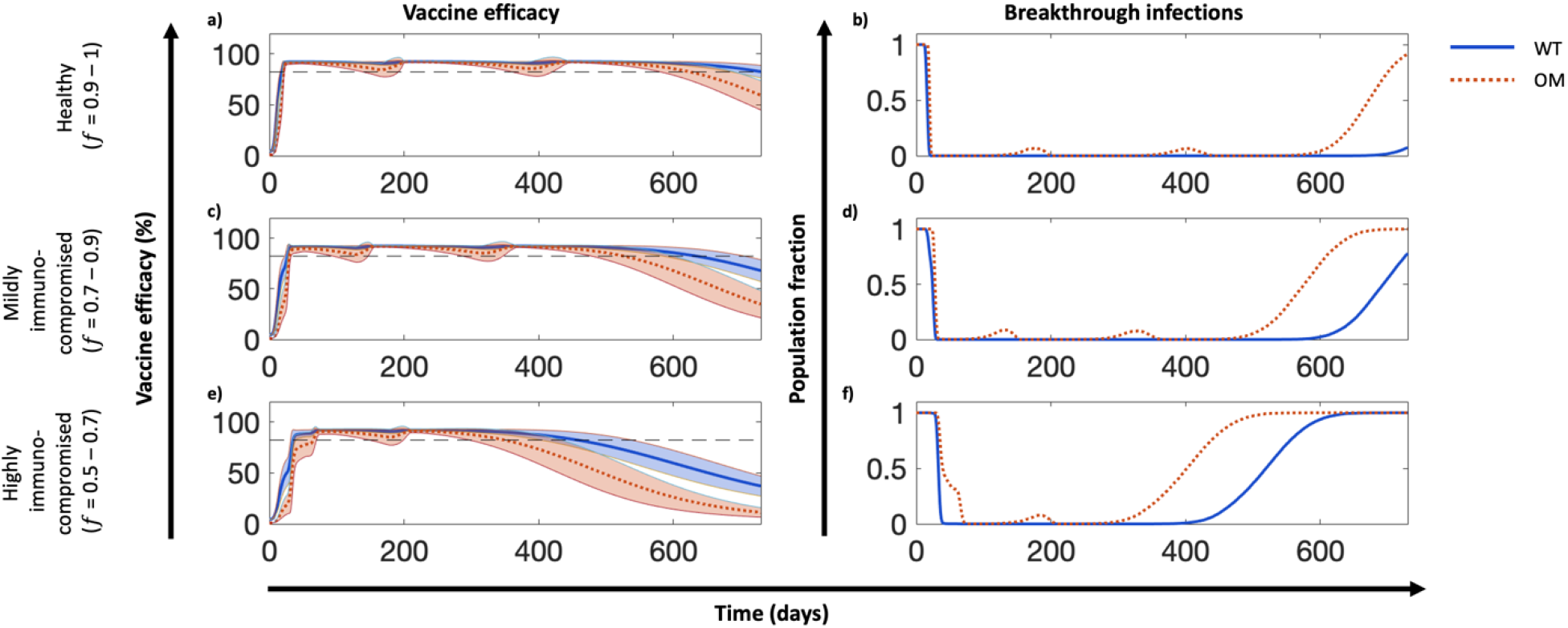
Testing model-predicted optimal dosing schedules. **a,c,e)** Vaccine efficacy and **b,d,f**) vulnerability to breakthrough infections due to wildtype strain (WT, solid blue line) and Omicron variant (OM, dotted orange line) of SARS-CoV-2 in **a,b**) healthy, **c,d**) mildly immunocompromised, and **e,f**) highly immunocompromised individuals. For each population sub-type, testing was done on 10,000 simulated individuals with unique *f* and dosing schedule values. Colored bands represent 90% prediction intervals.

Finally, the optimal dosing schedules identified above are summarized in **Figure 9** (green bands), with a comparison made to the CDC-recommended dosing schedules being currently implemented for the Pfizer-BioNTech vaccine (blue bands). The ongoing CDC guidelines for COVID-19 vaccination for healthy individuals (not moderately or severely immunocompromised and <50 years of age) include 3 doses with intervals of 3-8 weeks between the first and second dose (represented as 21 days) and 5 months between the second and third dose (represented as 140 days). The model-predicted schedule closely recapitulates the CDC guidelines with the inclusion of a fourth dose to prolong immunity for 385 days (>1 year). Although the model distinguishes between two immunocompromised cancer populations (mildly and highly), the CDC guidelines suggest a schedule of 4 doses for patients who are moderately or severely immunocompromised (with intervals of 21, 21, and 84 days, respectively). According to the model-predicted optimal dosing schedule, longer gaps between doses (or boosters) would not compromise the immunity of healthy and immunocompromised patients that could represent a solution to logistic constraints.

**Figure 9.**
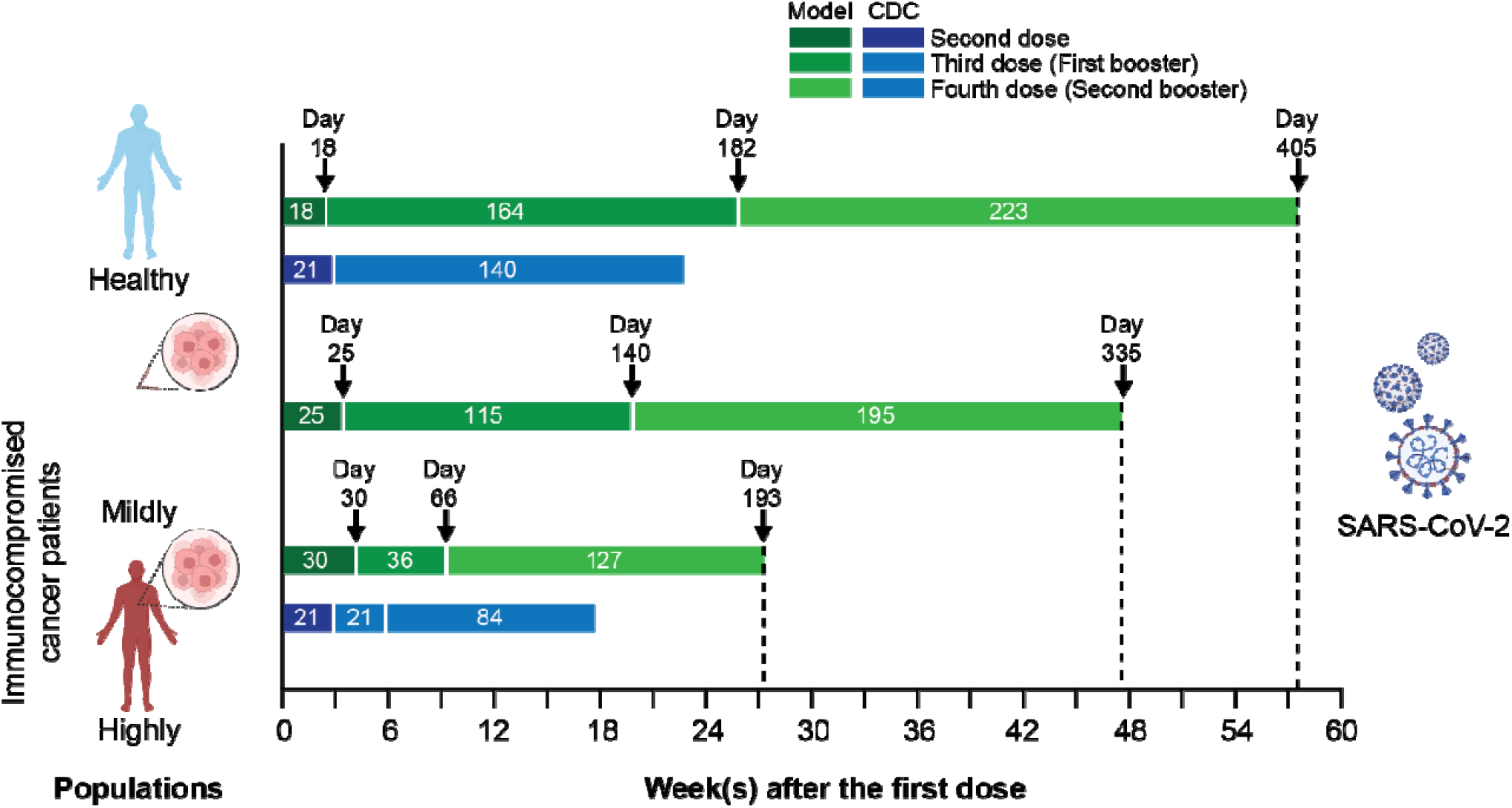
Model-predicted optimal dosing and CDC-recommended dosing schedules for the Pfizer-BioNTech vaccine in healthy and immunocompromised sub-populations. The ongoing CDC guidelines for dosing schedules are represented by the blue bands and those predicted by the model are shown in green.

## 4. Conclusions

In summary, we developed a mechanistic mathematical model of adaptive immune response to COVID-19 vaccines and viral infection in healthy and immunocompromised individuals. The model was formulated as a system of ODEs to account for key biological interactions leading to the development of antigen-induced humoral and cellular immunity. Following the calibration and validation of the model with published clinical data, numerical experiments were performed to study the effects of immune health status and vaccine dosing schedules on plasma antibody titers (a correlate of protection against infection), leading to the estimation of population vulnerability to breakthrough infections. Through simulations of virtual individuals, the model was then applied to identify optimal dosing schedules of the vaccines to minimize breakthrough infections in the population. Through our analysis, we highlighted critical waiting windows for immunocompromised individuals (25 days and 30 days after first dose for mildly and highly immunocompromised individuals, respectively) to ensure sufficient time for the development of immune recall responses and minimize vulnerability to breakthrough infections in their sub-populations. In the case of healthy individuals, while the optimal waiting period after first dose was found to be two and a half weeks, we proposed that it can be extended (without much compromise to protection) up to six weeks. Thereby, we make the case for longer waiting period between doses without compromising the immunity at the population scale. The presented model is based on generalized adaptive immune response to antigens and can thus be adapted to investigate different infections or vaccines, given appropriate data for model calibration. Through our proof-of-concept study, we have thus presented a novel approach to optimizing vaccine dosing schedules in case of future outbreaks. While the current model is thoroughly calibrated and validated, the mechanistic underpinnings of immunosuppression and innate immune response need to be considered in greater detail in future studies. Also, model adaptations relevant to the other types of COVID-19 vaccines may need to be considered as well. Lastly, our results also suggest the need for follow-up boosters (more frequently for immunocompromised subjects due to rapidly waning immunity) to ensure continued immunity against breakthrough infections and reinfections, especially given the emergence of novel VOCs.

## Data Availability

All data produced in the present study are available upon reasonable request to the authors.

https://www.nature.com/articles/s41586-020-2588-y

https://www.nejm.org/doi/10.1056/NEJMc2115596

https://www.esmoopen.com/article/S2059-7029(21)00236-2/fulltext

https://www.mdpi.com/2076-393X/9/10/1092

https://www.mdpi.com/2076-393X/10/6/876

## Author contributions

PD conceived and supervised the study. PD and CS curated the data, developed the mathematical model, and performed modeling analysis and simulations. PD, CS, ZW, JRR, SC, and VC designed numerical experiments. PD, CS, DIS, CM, JW, HDS, RP, and WA helped with interpretation of modeling results. All authors contributed to manuscript writing and editing.

## Acknowledgements

This study was conducted under the umbrella of the International Academic Affiliation Agreement between the Houston Methodist Academic Institute (Houston, TX, USA) and the University of Naples Federico II (Naples, Italy). The work was supported in part by the Cockrell Foundation (PD, VC), the National Institutes of Health (NIH) Grants 1R01CA253865 (ZW, VC), 1R01CA222007 (ZW, VC), and 1R01CA226537 (ZW, VC, RP and WA), Rutgers Cancer Institute of New Jersey (NCI Cancer Center Support Grant number P30CA072720), and by awards from the Levy-Longenbaugh Donor-Advised Fund (to RP and WA). The funders had no role in study design, data collection and analysis, decision to publish, or preparation of the manuscript. We thank Prof. Alan S. Perelson for his valuable criticism of our work. PD and CS also acknowledge Luca Messina and Maria J. Peláez for helpful scientific discussions, Rachael E. Whitehead for her contributions to the model schematic, and Life Science Editors for editing support.

