## Supplementary Information for "A modeling-based approach to optimize COVID-19 vaccine dosing schedules for improved protection"

**Antibody titer as a correlate of vaccine efficacy (or protection)**

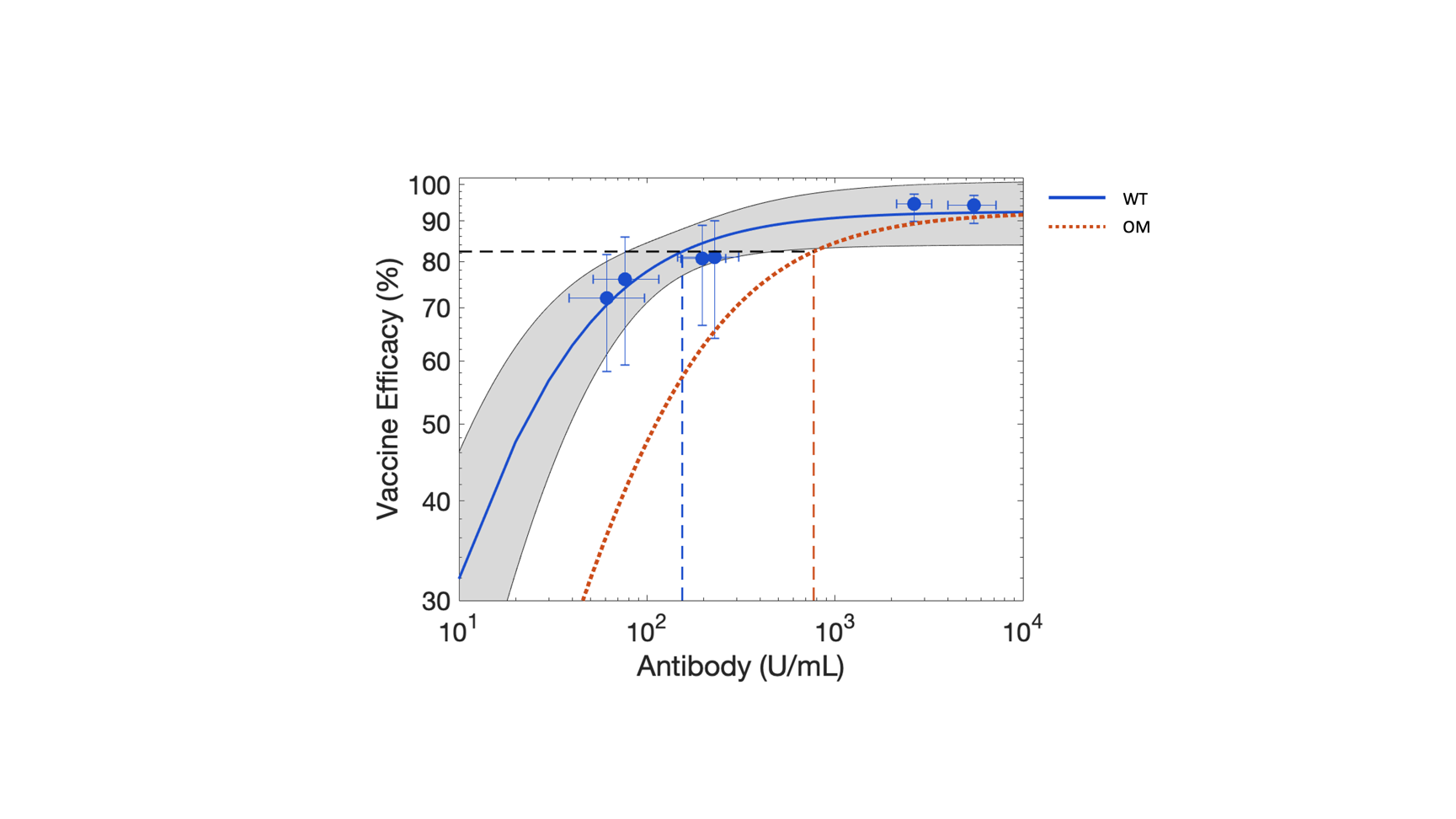

**Figure S1.** Least squares fitting of Michaelis-Menten function (Equation 18; solid blue curve) to clinical data (blue circles), obtained from Goldblatt et al. [1], for the effect of antibody (IgG) titer on vaccine efficacy against wild type (WT) strain of SARS-CoV-2. Gray band is the 95% confidence interval band. Dotted orange curve represents the corresponding vaccine efficacy prediction (based on Equation 19) for the Omicron (OM) strain of SARS-CoV-2. Error bars indicate 95% confidence intervals for either the antibody level (x-axis) or vaccine efficacy (y-axis). Dashed blue and orange lines denote antibody titer values of 154 U/mL and 770 U/mL, which corresponds to 82.3% vaccine efficacy (dashed black line) against WT and OM, respectively. Note: x- and y-axes are in log scale.

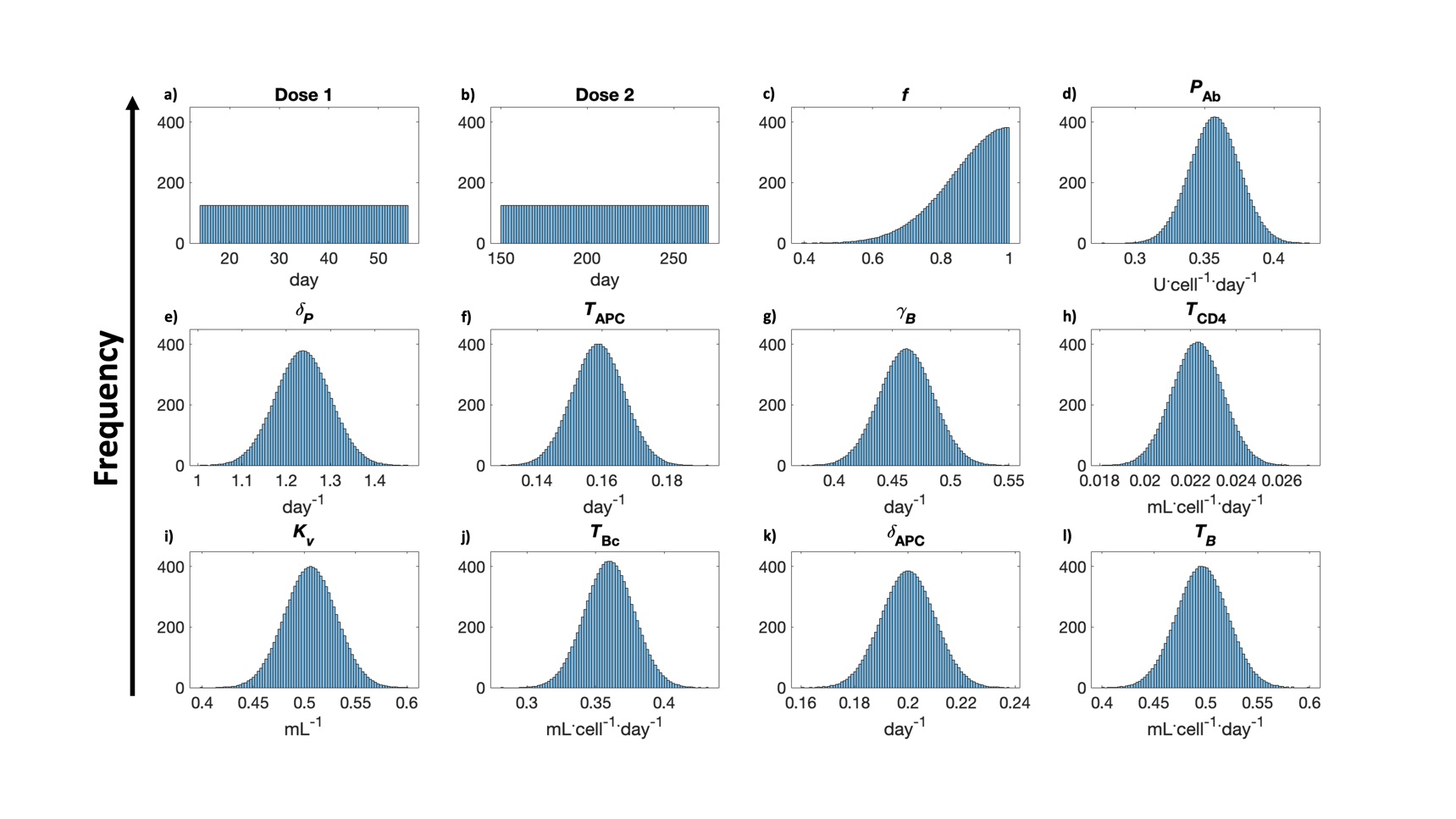
**Parameter distributions for virtual cohort generation**

**Figure S2.** Representative distributions (N = 10,000) of model parameters, identified from GSA as highly sensitive parameters, used for virtual clinical simulations.

**Pearson correlation analysis (model calibration)**

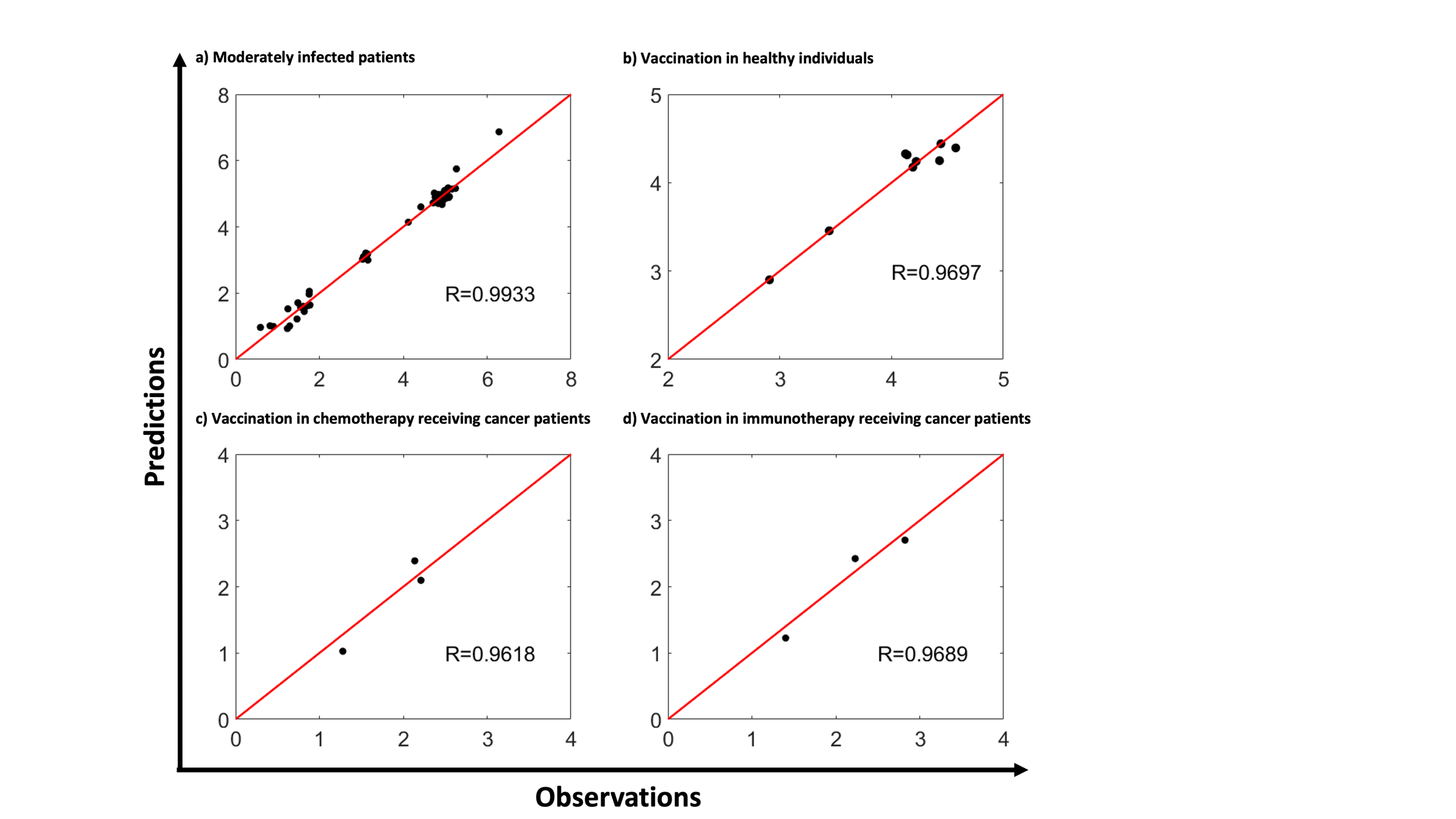

**Figure S3.** Pearson correlation analysis between model fits and experimental data for immune response dynamics (comprising one or more of the following variables: viral load, naïve and effector CD4+ and CD8+ T-cells, type-I and type-II interferon, IL-6, and neutralizing antibody (IgG)) during **a)** moderate SARS-CoV-2 infection, vaccination in **b**) healthy individuals, **c**) cancer patients receiving chemotherapy, and **d**) cancer patients receiving immunotherapy. R value represents Pearson correlation coefficient.

**Pearson Correlation analysis (model validation)**

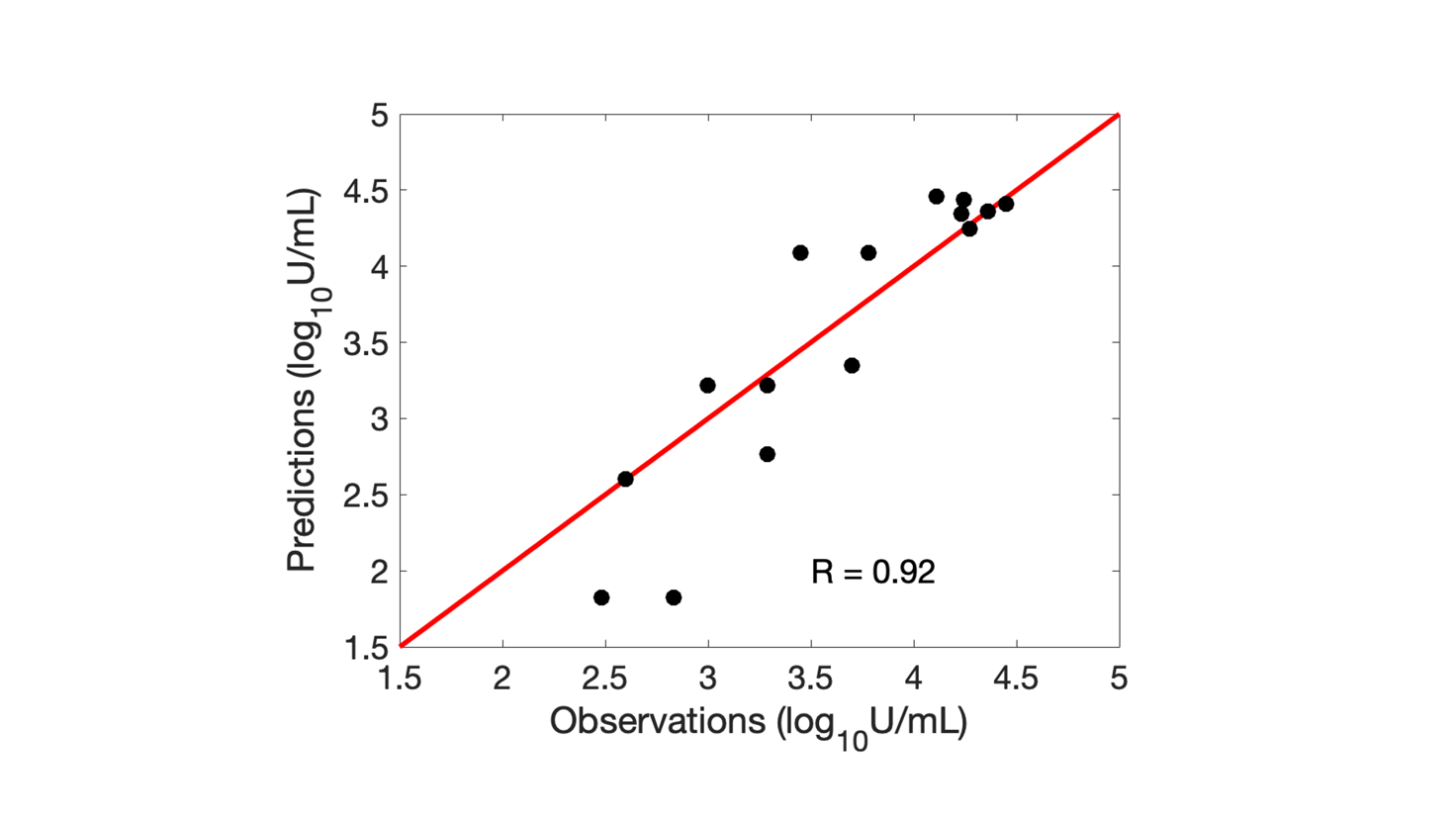

**Figure S4.** Pearson correlation analysis between model predictions and experimental data pooled from clinical studies on antibody (IgG) response dynamics following two doses of Moderna COVID-19 mRNA vaccine, and two and three doses of Pfizer-BioNTech COVID-19 mRNA vaccine. R value represents Pearson correlation coefficient.

**Local sensitivity analysis**

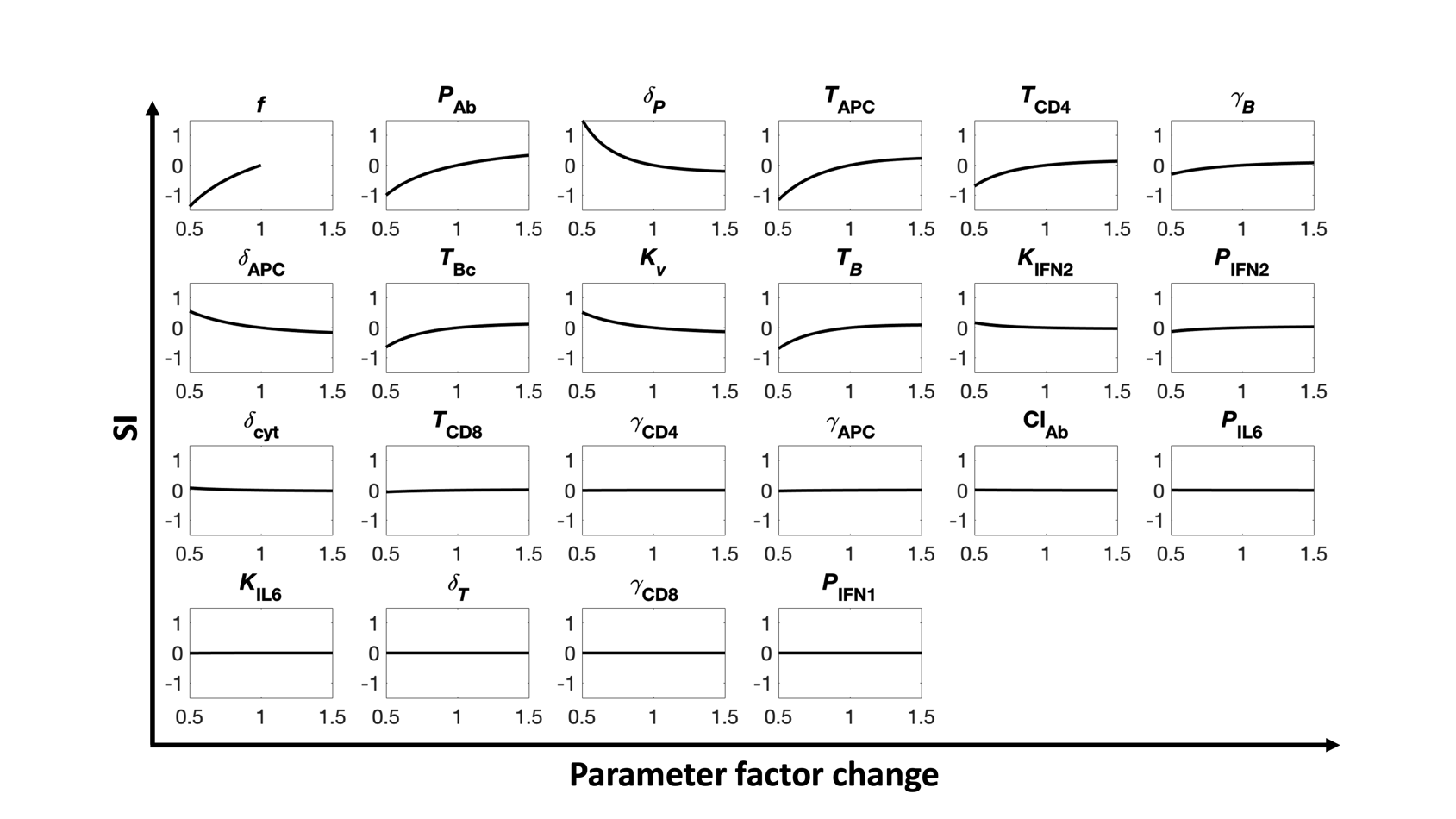

**Figure S5.** Local sensitivity analysis exhibiting correlation between parameter perturbation and sensitivity index. Parameters were perturbed linearly between $\pm$50% around the baseline value, except $f$ that was perturbed between –50% to baseline.
